# Multi-omics insights into the biological mechanisms underlying gene-by-lifestyle interactions with smoking and alcohol consumption detected by genome-wide trans-ancestry meta-analysis

**DOI:** 10.1101/2021.07.26.21261153

**Authors:** Timothy D. Majarian, Amy R. Bentley, Vincent Laville, Michael R. Brown, Daniel I. Chasman, L. Adrienne Cupples, Paul S. de Vries, Mary F. Feitosa, Nora Franceschini, W. James Gauderman, Daniel Levy, Alanna C. Morrison, Michael Province, Dabeeru C. Rao, Karen Schwander, Yun Ju Sung, Charles N. Rotimi, Hugues Aschard, C. Charles Gu, Alisa K. Manning, on behalf of the CHARGE Gene-Lifestyle Interactions Working Group

## Abstract

Gene-lifestyle interaction analyses have identified genetic variants whose effect on cardiovascular risk-raising traits is modified by alcohol consumption and smoking behavior. The biological mechanisms of these interactions remain largely unknown, but may involve epigenetic modification linked to perturbation of gene expression. Diverse, individual-level datasets including genotypes, methylation and gene expression conditional on lifestyle factors, are ideally suited to study this hypothesis, yet are often unavailable for large numbers of individuals. Summary-level data, such as effect sizes of genetic variants on a phenotype, present an opportunity for multi-omic study of the biological mechanisms underlying gene-lifestyle interactions. We propose a method that unifies disparate, publicly available summary datasets to build mechanistic hypotheses in models of smoking behavior and alcohol consumption with blood lipid levels and blood pressure measures. Of 897 observed genetic interactions, discovered through genome-wide analysis in diverse multi-ethnic cohorts, 48 were identified with lifestyle-related differentially methylated sites within close proximity and linked to target genes. Smoking behavior and blood lipids account for 37 and 28 of these signals respectively. Five genes also showed differential expression conditional on lifestyle factors within these loci with mechanisms supported in the literature. Our analysis demonstrates the utility of summary data in characterizing observed gene-lifestyle interactions and prioritizes genetic loci for experimental follow up related to blood lipids, blood pressure, and cigarette smoking. We show concordance between multiple trait-or exposure-related associations from diverse assays, driving hypothesis generation for better understanding gene-lifestyle interactions.

## Introduction

Lifestyle and environmental exposures have been shown to modify the underlying genetic risks of health outcomes(1-5). Recent large scale studies(6-10) have sought to characterize these interactions through genome-wide interaction analyses among tens of thousands of people from diverse ancestral backgrounds, implicating genetic loci for which the genetic effect on a phenotype of interest is modified by either alcohol consumption or cigarette smoking. The mechanisms through which these interactions act remain largely unexplained. A plausible explanation for these interactions is a combination of genetic and epigenetic mechanisms of action, such as differential methylation of important gene regulatory regions by genotype. Evaluating this possibility requires harmonizing disparate data types and jointly analyzing with genetic variants in models that account for interaction effects.

The harmonization of multi-omics and genetic data presents a challenge. With current genetic research paradigms, obtaining and matching multi-modal data for an individual is often not possible given large, complex hurdles in place, necessary to protect individual privacy. Summary-level metrics, such as effect sizes of genetic variants on a phenotype, are often more easily obtainable through consortia data-sharing agreements and shielded from exposing the identity or personal information of the study participants. Therefore, an analysis of association data that relies on summary results alone is particularly valuable in this context.

To this end, we propose a gene-centric prioritization method that leverages multiple datasets of summary results (genetic, epigenetic, and transcriptomic) to better understand and evaluate the possible mechanisms through which gene-environment interactions have been observed. Here, we use data from the CHARGE Consortium’s Gene-Lifestyle Interactions Working Group(11), published in five recent papers(6-10) together with differential methylation and gene expression data from the literature(12-19) to generate mechanistic hypotheses for gene-environment interactions centered on gene regulation. We demonstrate that by considering diverse epigenetic associations at the same genetic loci and linking multiple forms of evidence for common variants to a gene, gene-environment interactions may be more fully characterized and prioritized for further mechanistic studies with the goal of providing biological and translational insights.

## Results

We used variant-level summary statistics from genome-wide gene-lifestyle interaction (GLI) analyses of cigarette smoking habits (current or ever smoking) and alcohol consumption (current or heavy vs. light drinking) with lipid traits (high-density lipoprotein cholesterol [HDL], low-density lipoprotein cholesterol [LDL], and triglycerides [TG]) and blood pressure traits (systolic blood pressure [SBP], diastolic BP [DBP], mean arterial pressure [MAP], and pulse pressure [PP]) (**Table 1; Supplemental Table 1**). The generation(6-10) and harmonization(20) of these summary statistics has been previously described, resulting in 140 sets of results in four ancestry groups and one trans-ancestry meta-analysis (**Supplementary Figure 1**). To restrict our efforts to regions in the genome most likely to have a GLI effect on the trait, we considered genetic variants with interaction p-value less than 5×10^−5^ which resulted in 897 variants (**Figure 1**). Of these variants, 682 were seen in smoking behavior interaction models, 511 were seen in models focusing on blood pressure traits, and 674 were observed in the analyses in the African ancestry subset (**Figure 2**).

**Table 1:** Datasets used in locus characterization and prioritization.

| Trait/exposure | Type | Source | Analysis | Data Extracted |
| --- | --- | --- | --- | --- |
| Blood Pressure/Smoking | Genetic Association | Sung *et al.* (2018)<br>Sung *et al.* (2019) | GLI | 464 SNPs |
| Lipids/Smoking | Genetic Association | Bentley *et al.* (2019) | GLI | 218 SNPs |
| Blood Pressure/Alcohol | Genetic Association | Feitosa *et al.* (2018) | GLI | 47 SNPs |
| Lipids/Alcohol | Genetic Association | de Vries *et al.* (2019) | GLI | 168 SNPs |
| Blood Pressure & Lipids/Smoking & Alcohol | Genetic Association | Laville *et al.* (2021*) | GLI | Harmonized summary data of above GLI projects |
| Lipids | Methylation | Dekkers *et al.* (2016) | DMe | 28 DNAm* |
| Blood pressure | Methylation | Richard *et al.* (2018) | DMe, eQTM | 126 DNAm, 13 eQTM* |
| Smoking | Methylation | Johannes *et al.* (2016) | DMe, eQTM | 2,623 DNAm, 1,430 eQTM* |
| Alcohol | Methylation | Liu *et al.* (2018) | DMe, eQTM | 328 DNAm, 14,160 eQTM* |
| Smoking | Gene Expression | Parker *et al.* (2017) | DExpr | 171 genes* |
| Smoking | Gene Expression | Huan *et al.* (2016) | DExpr | 1,270 genes* |
| Smoking | Gene Expression | Nikodemova *et al.* (2018) | DExpr | 25 genes* |
| - | Gene Expression | Lonsdale *et al.* (2013) | eQTL | Genes associated with GLI SNPs from all tissues |

\* Analyses performed in whole blood

**Figure 1.**
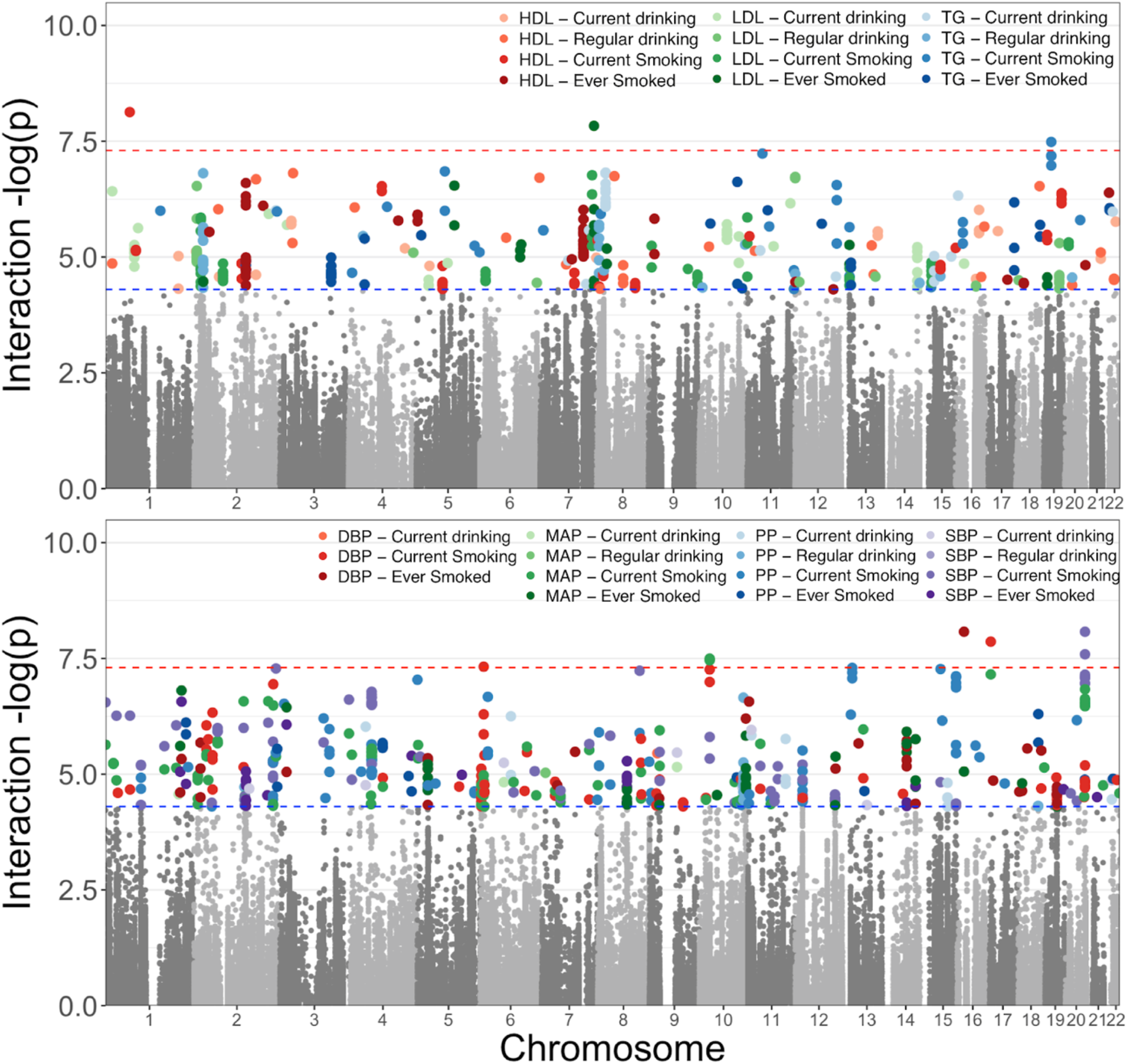
Selection of GLI Associations for Evaluation; a total of 897 variants were selected from meta-analyses of four ancestral groups and one trans-ancestry group of the variant interactions with 2 smoking and 2 alcohol exposure variables on 3 lipid measures and 4 blood pressure traits. Shown are the most statistically significant associations across the 5 meta-analyses for each of the phenotype-exposure combinations. Results separated by ancestry group available in **Supplemental Figure 1**.

**Figure 2.**
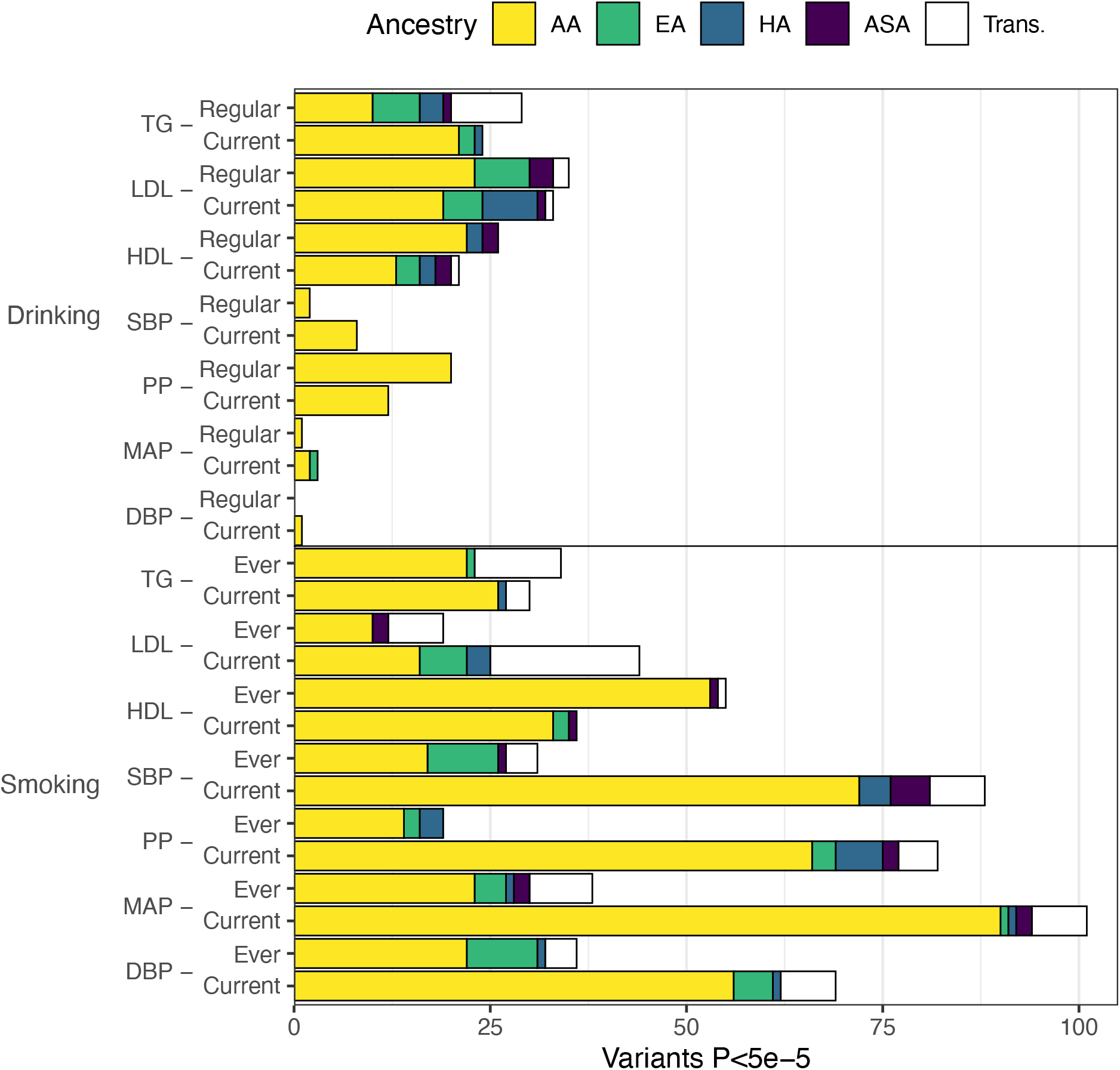
Distribution of GLI Associations across Phenotypes and Exposures; the total number of associated genetic variants discovered in each trait-exposure-ancestry group is displayed. Associations in models utilizing alcohol-related exposures are shown at top and represent the minority of observed associations. Alcohol-related associates were dominated by analyses studying blood lipids. Associations related to cigarette smoking exposure are shown at bottom with the majority of observations among analyses of blood pressure measures.

In an effort to build a more complete picture of gene regulatory mechanisms that could be contributing to the interaction signal, we found loci in which nominal GLI effects and significant epigenetic effects were observed (**Figure 3A**). We used a variety of association data from cohort studies, including differential methylation analysis (DMe analysis; see **Methods**) with both clinical outcomes (lipid and blood pressure traits) and lifestyle outcomes (smoking behaviors and alcohol consumption habits), and differential gene expression analysis (DExpr analysis; see **Methods**) conditional on smoking behaviors (**Table 1**). For the traits and exposures considered in the genetic association analysis, we compiled a list of loci with significantly differentially expressed genes or differentially methylated methylation sites (DNAm), extending the borders of the gene (defined by exon boundaries) or site by 500 KB. This intersection of genetic, epigenetic, and transcriptomic data resulted in 833 unique genes (**Figure 3)** multiple forms of evidence for association with the trait and exposure.

**Figure 3.**
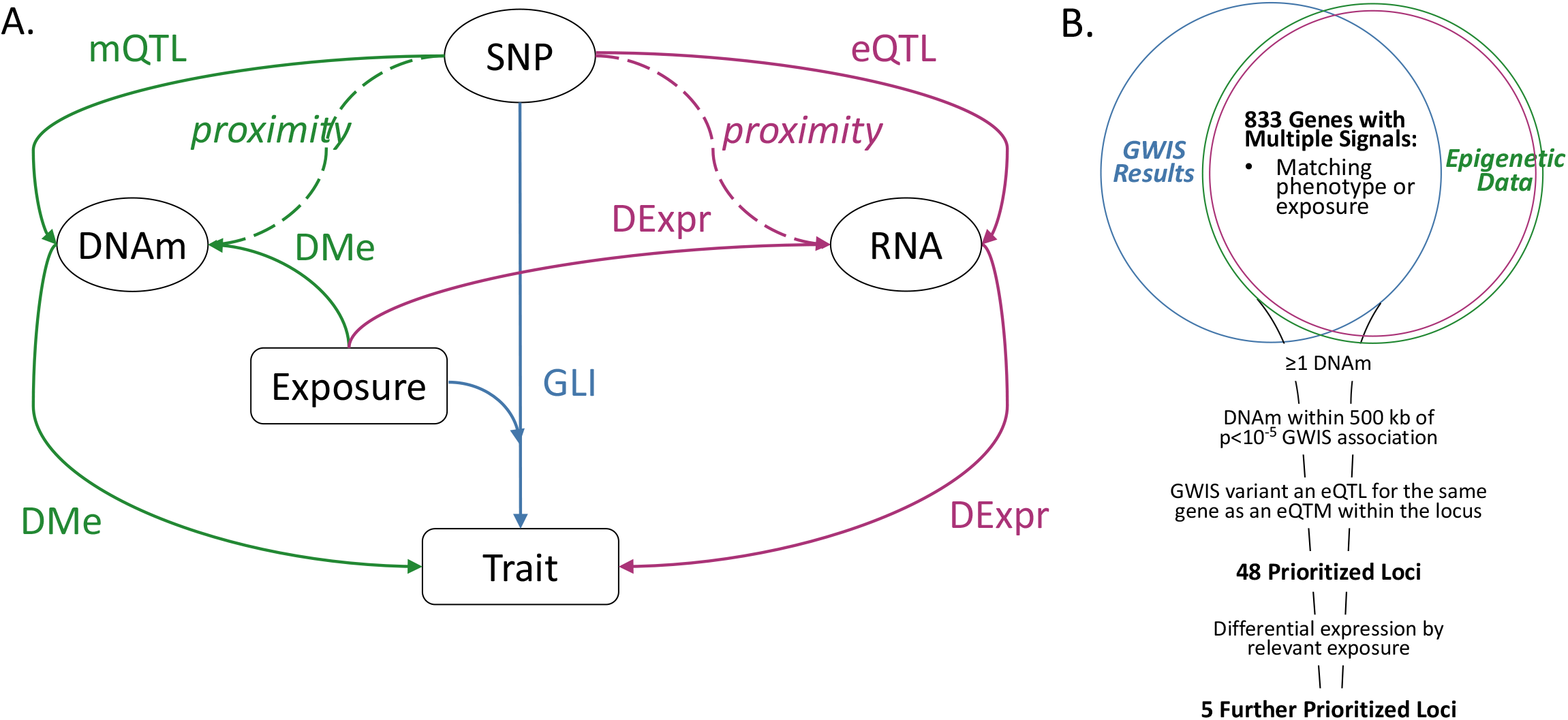
(A) Illustration of Possible Mechanisms Underlying Interaction Signals; rectangles represent study traits or measured lifestyle exposures; ovals represent 3 types of molecular risk factors: SNP genotype, RNA expression, and DNA methylation. Interactions are depicted by the “SNP-Exposure-Trait” path (GLI), with possible underlying molecular mechanisms shown along the sides for the transcriptomic (RNA expression) and epigenomic level (DNAm). The molecular effects are represented by solid arrows: expression QTL effect of a SNP allele on RNA expression levels (eQTL analysis) or methylation (mQTL analysis); differential methylation of a CpG site by trait or exposure (DMe analysis); and differential expression by trait or exposure (DExpr analysis). Dashed lines indicate physical proximity between elements. (B) Prioritization of GLI Loci; a panel of GLI loci from multiple genome-wide interaction studies were intersected with publicly-available epigenomic and transcriptomic data for the relevant variants, lifestyle exposures, and traits. The resulting data were then prioritized using the listed criteria to yield 48 loci with multiple sources of molecular evidence underlying the interaction and 5 loci with differential expression by the exposure of interest.

To further analyze these genic regions, we required loci to contain at least one DNAm site associated with expression of a gene (eQTM analysis; see **Methods**) and at least one of the GLI variants to be associated with the expression of the same gene in at least one tissue (eQTL analysis; see **Methods**). Using these criteria, we prioritized 48 signals at 36 loci. Of these, 37 were linked to smoking behavior and 28 were linked to lipid outcomes (**Table 2**). The majority of prioritized signals (N=30) were derived from analyses in individuals of African ancestry, 26 of which were related to smoking behavior. These signals were evenly distributed between lipid and blood pressure traits. Our final prioritization step matched the lifestyle exposure from the GLI effect at the locus with significant genes from the DExpr analysis of the same exposure. At the 48 prioritized signals, no genes showed differential expression with any alcohol exposure while five genes showed differential expression with a smoking exposure: *GCNT4, PTPRZ1, SYN2, ALDH2, TMEM116* (**Figure 3B**; **Supplemental Figures 2-8**).

**Table 2:** Genes prioritized by GLI association signal, differential methylation, and presence of eQTLs.

| Exposure | Gene Target | eQTL Tissue(s) | Number of GLI variants |
| --- | --- | --- | --- |
| *African Ancestry HDL* |  |  |  |
| Curr Smk | *POLK* | Heart Left Ventricle, Pancreas, Thyroid<sup>1</sup> | 5 |
| Curr Smk | *GCNT4*<sup>2</sup> | Artery Aorta, Testis | 4 |
| Curr Smk | *KLK8* | Skin Sun Exposed Lower leg | 4 |
| Curr Smk | *ANGPT1* | Thyroid | 3 |
| Curr Smk | *CARM1*<sup>3</sup> | Muscle Skeletal | 1 |
| Curr Smk | *LIPC* | Liver, Pancreas | 1 |
| Curr Smk | *YIPF2*<sup>3</sup> | Lung | 1 |
| Ev Smk | *PTPRZ1*<sup>2,3</sup> | Artery Aorta | 21 |
| Ev Smk | *DNAH7* | Adipose Visceral Omentum, Pancreas, Thyroid<sup>1</sup> | 1 |
| *African Ancestry LDL* |  |  |  |
| Ev Smk | *CRYGN* | Colon Sigmoid, Nerve Tibial | 1 |
| Ev Smk | *FRK* | Muscle Skeletal | 1 |
| Ev Smk | *WDR86* | Nerve Tibial | 1 |
| *African Ancestry TG* |  |  |  |
| Curr Drnk | *BAHD1* | Colon Sigmoid, Stomach, Thyroid<sup>1</sup> | 2 |
| Curr Drnk | *DNAJC17* | Esophagus Mucosa | 2 |
| Ev Smk | *TRIO* | Ovary | 1 |
| *African Ancestry DBP* |  |  |  |
| Ev Smk | *FNTB* | Muscle Skeletal, Skin Sun Exposed Lower leg | 3 |
| Ev Smk | *RAB15* | Adipose Subcutaneous, Nerve Tibial, Thyroid<sup>1</sup> | 3 |
| *African Ancestry MAP* |  |  |  |
| Curr Smk | *CRTAC1* | Adipose Visceral Omentum, Artery Tibial | 1 |
| Curr Smk | *SULT4A1* | Colon Sigmoid | 1 |
| Ev Smk | *FNTB* | Muscle Skeletal, Skin Sun Exposed Lower leg | 3 |
| Ev Smk | *RAB15* | Adipose Subcutaneous, Nerve Tibial, Thyroid<sup>1</sup> | 3 |
| Ev Smk | *KCNK13* | Brain Hypothalamus | 1 |
| *African Ancestry PP* |  |  |  |
| Curr Drnk | *MPI* | Artery Aorta, Lung, Whole Blood<sup>1</sup> | 4 |
| Curr Drnk | *SCAMP2* | Esophagus Mucosa, Pituitary, Whole Blood<sup>1</sup> | 4 |
| Curr Smk | *SYN2*<sup>2</sup> | Artery Aorta | 1 |
| Ev Smk | *JMJD4* | Nerve Tibial | 2 |
| *African Ancestry SBP* |  |  |  |
| Curr Smk | *CRTAC1* | Adipose Visceral Omentum, Artery Tibial | 1 |
| Ev Smk | *FNTB* | Muscle Skeletal, Skin Sun Exposed Lower leg | 2 |
| Ev Smk | *RAB15* | Adipose Subcutaneous, Nerve Tibial, Thyroid<sup>1</sup> | 2 |
| Ev Smk | *JMJD4* | Nerve Tibial | 1 |
| *Asian Ancestry HDL* |  |  |  |
| Ev Smk | *KCTD10*<sup>3</sup> | Artery Tibial, Skin Sun Exposed Lower leg | 1 |
| Ev Smk | *MYO1H*<sup>3</sup> | Lung | 1 |
| Ev Smk | *UBE3B*<sup>3</sup> | Colon Transverse, Skin Sun Exposed Lower leg | 1 |
| *Asian Ancestry LDL* |  |  |  |
| Drnk habits | *CPNE2* | Cells Transformed fibroblasts | 3 |
| Ev Smk | *KANK2*<sup>3</sup> | Adipose Subcutaneous, Lung, Thyroid<sup>1</sup> | 2 |
| *Asian Ancestry TG* |  |  |  |
| Drnk habits | *LIPC*<sup>3</sup> | Thyroid | 1 |
| *Asian Ancestry MAP* |  |  |  |
| Ev Smk | *ALDH2*<sup>2,3</sup> | Esophagus Mucosa, Lung, Nerve Tibial, Thyroid<sup>1</sup> | 2 |
| Ev Smk | *TMEM116*<sup>2,3</sup> | Artery Tibial, Heart Atrial Appendage, Whole Blood<sup>1</sup> | 1 |
| *Asian Ancestry SBP* |  |  |  |
| Curr Smk | *ALDH2*<sup>2,3</sup> | Esophagus Mucosa, Brain Cortex, Thyroid<sup>1</sup> | 3 |
| Ev Smk | *ALDH2*<sup>2,3</sup> | Esophagus Mucosa, Skin Sun Exposed Lower leg | 1 |
| *European Ancestry LDL* |  |  |  |
| Drnk habits | *APOC1*<sup>3</sup> | Esophagus Mucosa | 1 |
| *European Ancestry TG* |  |  |  |
| Drnk habits | *KRTCAP3*<sup>3</sup> | Adrenal Gland, Thyroid, Whole Blood<sup>1</sup> | 2 |
| Drnk habits | *PPM1G*<sup>3</sup> | Adipose Subcutaneous, Stomach, Thyroid<sup>1</sup> | 2 |
| *Hispanic Ancestry PP* |  |  |  |
| Curr Smk | *STIM1* | Lung, Thyroid, Whole Blood | 2 |
| *Trans Ancestry LDL* |  |  |  |
| Ev Smk | *CRYGN* | Colon Sigmoid, Nerve Tibial | 1 |
| Ev Smk | *WDR86* | Nerve Tibial | 1 |
| *Trans Ancestry TG* |  |  |  |
| Drnk habits | *KRTCAP3*<sup>3</sup> | Adrenal Gland, Thyroid, Whole Blood<sup>1</sup> | 1 |
| Drnk habits | *PPM1G*<sup>3</sup> | Esophagus Muscularis, Muscle Skeletal | 1 |

<sup>1</sup>An eQTL in multiple tissues: the most relevant 3 are shown (full results available in **Supplemental Table 2**); <sup>2</sup>Differential Expression by Exposure; <sup>3</sup>Loci with 2df GLI association; Abbreviations: Current Smoking (Curr Smk), Ever Smoking (Ev Smk), Drinking Habits (Drnk habits)

At the *GCNT4* locus, the common A allele at the rs3761743 variant was associated with decreased HDL levels in the ‘current smokers’ exposure group (P=6.9×10^−4^) in an African-ancestry subset of cohorts, an effect that was attenuated in the non-smoker exposure group (Interaction P=1.5×10^−5^). *GCNT4* is a glycosyltransferase expressed primarily in the thymus(21) but rs3761743 was associated with decreased expression of GCNT4 in the aorta (P=3.5×10^−6^) and increased expression in testis (P=6.7×10^−14^) (**Supplemental Table 3, Supplemental Figure 2**). Notably, *GCNT4* is upregulated among smokers(18) (P=9.8×10^−5^) while methylation of the DNAm site cg21158503 was decreased with smoking exposure(14) (P=6.6×10^−6^).

In the analysis of lipids and the ‘ever-smoking’ exposure in the African ancestry subset of cohorts, the association between the rs77810251 variant at the *PTPRZ1* locus and HDL levels was found to differ between exposure strata (Interaction P=9.5×10^−7^), with a positive association of the minor A allele in the ‘ever-smoking’ exposure group and no association among the ‘never-smoking’ exposure group(6) (**Figure 4A, Supplemental Table 4, Supplemental Figure 3**). rs77810251 is an eQTL for *PTPRZ1* in aorta tissue, with the A allele associated with decreased gene expression. A DNAm site within the locus, cg00826384, shows increased methylation among smokers(14). *PTPRZ1* was shown to be downregulated in nicotine-treated cells(22). These generate a potential hypothesis of rs77810251 (and other associated variants in this locus) leading to decreased expression of *PTPRZ1*, which may cause an increase in HDL (through unknown mechanisms). Smoking, which is associated with increased methylation of *PTPRZ1*, may perturb the *PTPRZ1*-HDL pathway, and abolish the association between rs77810251 and HDL. *PTPRZ1* is a protein tyrosine phosphatase receptor, which is constitutively active and is inactivated through binding with heparin-binding growth factors pleiotrophin and midkine. Its inactivation leads to increased tyrosine phosphorylation of target genes. This gene has a broad spectrum of substrates that may mediate multiple pathways(23). Of interest, both *PTPRZ1* and *LRP6*, a member of the well-established lipids signaling family of LDL-receptor related proteins, are regulated through binding with midkine(24), suggesting a potential connection of this locus with a lipids pathway.

**Figure 4.**
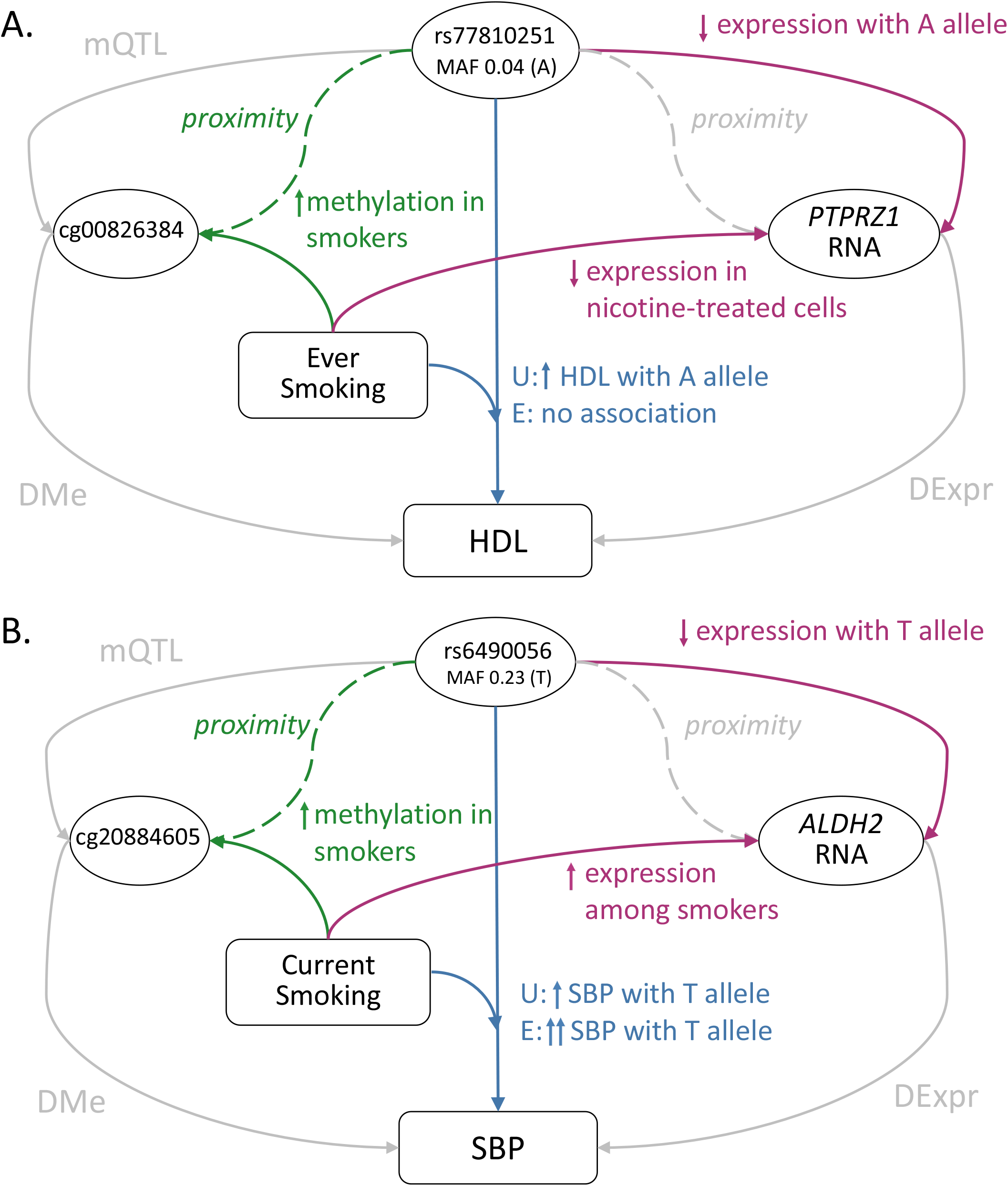
(A) Summary of evidence at *PTPRZ1* GLI Locus; the GLI variant rs77810251 in *PTPRZ1* is also an eQTL with downregulated expression in aortic tissue, and was in close proximity to the DNAm site cg00826384, which shows elevated methylation among smokers (see text for further details; pathways without evidence for this association have been grayed out). (B) Summary of evidence at *ALDH2* GLI Locus; the GLI variant rs6490056 in *ALDH2* is also an eQTL with downregulated expression in multiple tissues, and was in close proximity to the DNAm site cg20884605, which shows elevated methylation among smokers (see text for further details; pathways without evidence for this association have been grayed out).

*SYN2* was prioritized from the PP and ‘current smoking’ GLI analysis among the African ancestry subset of cohorts. A rare T allele at a single variant in the locus, rs4135300, was associated with PP (P=8.72×10^−7^) with a positive effect in the ‘non-current smoking’ exposure group and a negative effect in the ‘current smoking’ exposure group (Interaction P=3×10^−7^; **Supplemental Table 5**). rs4135300 is an eQTL for *SYN2* in aorta tissue, with the T allele associated with decreased expression levels (**Supplemental Figure 4**). The DNAm site identified within the locus, cg10245988, has increased methylation on average among smokers(14). Expression of *SYN2* is increased in human neuroblastoma cells treated with nicotine. *SYN2* has previously been associated with schizophrenia(25).

The gene *ALDH2* was prioritized based on multiple analyses of smoking and blood pressure among the Asian ancestry subset of cohorts, specifically, the SBP with ‘current smoking’ (**Figure 4B; Supplementary Table 7; Supplementary Figure 6**), SBP with ‘ever smoking’ (**Supplementary Table 8; Supplementary Figure 7**), and MAP with ‘ever smoking’ **Supplementary Table 6; Supplementary Figure 5**) GLI analyses. As a representative example, the common T allele of rs6490056 was associated with increased SBP levels (P=1.9×10^−10^) with a larger effect in the ‘current smoking’ exposure strata (Interaction P=2.6×10^−5^). This variant and other associated variants in this locus are eQTLs for *ALDH2* in multiple tissues, including in the lungs and esophagus. *ALDH2* is a part of the major oxidative pathway for alcohol metabolism, and an Asian ancestry-specific variant (rs671) in this gene is well known for causing acetaldehyde accumulation with alcohol intake, leading to unpleasant side effects(26). Acetaldehyde and other toxic aldehydes are also components of tobacco smoke(27), and decreased ALDH2 activity leads to increased reactive aldehyde species and oxidative stress(28). In individuals deficient in ALDH2 activity, smoking amplifies risk of oxidative stress-related conditions(28). Importantly, oxidative stress is a known contributor to worsening blood pressure(29, 30). Based on these data, it appears possible that rs6490056 reduces expression of *ALDH2*, raising oxidative stress, and causing a concomitant increase in blood pressure. Under the additional burden of oxidative stress introduced by smoking, an even stronger effect on blood pressure traits may be observed through this locus.

*TMEM116* is a transmembrane protein, expressed in nearly all measured tissues(31). Two variants at this locus (rs6490056, rs10849962) were associated in MAP with ‘ever smoking’ GLI analyses among the Asian ancestry subset of cohorts (**Supplemental Table 9, Supplemental Figure 8**). The common T allele of rs6490056 was associated with increased MAP measures (P=5.7×10^−11^) with a larger effect in ‘ever smoking’ exposure group (Interaction P=3.3×10^−5^). These variants were significant eQTLs with the T allele of rs6490056 associated with lower *TMEM116* expression levels in atrial and adipose tissues and the A allele of rs10849962 associated with higher *TMEM116* expression levels in a variety of tissues, including esophageal tissues, heart tissue, adipose, and whole blood. *TMEM116* is upregulated among smokers with corresponding demethylation of a nearby DNAm site, cg08528204(13).

## Discussion

With the emergence of gene expression and methylation data in human cohorts, epidemiological study designs can now expand to epigenomic and transcriptomic hypotheses with the potential of understanding the regulation of pathways relevant to clinical phenotypes. These new insights may help explain gene-lifestyle interactions not typically captured by GWAS. The establishment of CHARGE’s Gene Lifestyle Interactions Working Group resulted in a number of large-scale, trans-ancestry evaluations of gene-by-environment interactions(6-10), with statistical evidence for interactions observed, yet the underlying mechanisms remained to be elucidated. In this analysis, we leveraged publicly available epigenomic and transcriptomic association data in order to identify a subset of interactions from these analyses among which multiple forms of evidence point toward potential mechanistic explanations. We identified 833 genes for which evidence of a nearby statistical interaction can be supported with functional data from at least one source, with 48 of these having multiple sources of functional support.

A striking finding from this work is the large proportion of interactions that are observed among African ancestry meta-analyses compared to other ancestries or to Trans-ancestry analyses. This phenomenon was observed among both the full number of interactions identified (79.7%) as well as the prioritized loci (62.5%), and 3 of the 5 further prioritized loci were from African ancestry meta-analyses. While this disproportion was evenly distributed among traits considered, it was not evenly distributed among exposures, with 26 of 30 of the prioritized loci based on meta-analyses of those of African ancestry were related to smoking exposure. For these 30 loci, the lead associated variant was African ancestry-specific (only present in 1KG AFR populations) for only 2, while for most the lead variants were available in all ancestries, but not associated in the meta-analyses of other ancestries. These results suggest that the source of this observation relates to a smoking exposure-related difference by ancestry.

Although we did not have the data for a detailed evaluation of smoking patterns (ex. type of cigarette, length of smoking history, age at first cigarette) by ancestry in our studies, there are pronounced differences in smoking patterns across ancestry groups in the US(32). Notably, there is a marked difference in type of preferred cigarette, as shown in data from the National Survey on Drug Use and Health, which is designed to be representative of the US population: 88% of African Americans smokers vs. 26% of non-Hispanic white smokers used menthol cigarettes(33). These differences in preference may stem from differences in bitter taste perception by ancestry, with menthol cigarettes more palatable to those with stronger bitter taste perception as the menthol flavor additive masks the bitterness of nicotine. An African ancestry-specific genetic locus, *MRGPRX4*, associated with a 5-to 8-fold increased odds of menthol cigarette smoking was recently identified(34), although only a small minority of African Americans carry this variant, suggesting more opportunities for novel explanation of menthol preference in African Americans.

Menthol cigarettes have long been targeted by the public health community based on evidence that they facilitate deeper smoke inhalation by decreasing nicotine-induced irritation^31^ (35). This deeper inhalation leads to a subsequent higher absorption of the myriad harmful components within cigarette smoke(32, 36). Consistent with this observation, ancestry differences in smoking-related metabolites and carcinogens have been reported(37-40), and different levels of these compounds may underlie the observed differences by ancestry in genetic interactions upon smoking exposure. Additionally, there is some evidence for greater systemic oxidative stress(41-44) and, relatedly, inflammation(45-48) among Americans of African vs. European ancestry. Exposure to cigarette smoke, a rich source of oxidants, on a background of elevated oxidative stress and inflammation may provoke a greater response among these individuals, manifesting as an interaction with smoking that differs by ancestry.

One key motivation for conducting GLI analyses is the relative ease of practical translation, as results suggest a modifiable risk, i.e. individuals with a certain genotype might reduce exposures associated with exacerbated risk. Confidence in such clinical translations, however, requires evidence beyond statistical findings of interactions. In our efforts to map existing functional information to loci of interest, we identified several areas where improvements in available data might facilitate stronger inferences. For instance, there are limited data to evaluate differential expression by alcohol exposure, making it difficult to further investigate the loci identified in these interaction analyses. Additionally, more tissue-specific data would be useful. Specific tissue types are of greater interest for each phenotype (e.g. liver for lipids) and for each exposure (e.g. lung for smoking); reliance on whole blood, which is the most available, will limit our understanding of the underlying biology. Data linking methylation to gene expression is also limited. Although it was beyond the study design of this project, it would be useful to collect individual level data to better elucidate these loci. Importantly, while some of the patterns observed in our data fit with expectations (e.g. the direction of RNA expression and DNA methylation for *GCNT4, PTPRZ1, TMEM116*), some did not, and more complete data is needed to flesh out these associations into a real understanding of the underlying mechanism. Our findings of interactions that differed by ancestry highlight the need for functional data from samples of diverse ancestries.

A strength of these analyses is the underlying interaction data. These data are drawn from discovery data on up to 133,805 individuals, an important observation given the statistical power needed to detect interactions. Additionally, the CHARGE GLI Working Group went to great effort to include studies of diverse ancestries, such that relatively large proportions of ancestry groups, such as African ancestry, that are underrepresented in genomic research, were achieved. Given the preponderance of African ancestry-identified associations among our results, this inclusion was of key significance. This work could have been improved with omics data derived from individuals of diverse ancestries to explore associations that differed by ancestry. The exposure data considered in these analyses was represented using binary variables in order to maximize sample sizes for detection of interactions, although the true effects of exposures are certain to be more complex, with variations with timing and dose of exposure. Similarly, the exposures and phenotypes we selected for these analyses were relatively straightforward to measure, however, a wide range of exposures may be involved in gene-lifestyle interactions on a wide range of phenotypes, and it is unknown whether these findings are representative of gene-lifestyle interactions in general. Also, additional experimental data will be necessary to confirm biological pathways suggested by these findings in order to advance the evidence from this work towards clinical translation.

In summary, this work provides preliminary evidence from publicly available transcriptomics and epigenomics data to support potential biological mechanisms underlying gene-lifestyle interactions identified through statistical evidence. These data suggest how GLI may occur, motivating future studies that include individual-level epigenomic and transcriptomic data, other environmental exposures and outcomes, and more complex characterization of exposure. This work also highlights the importance of including individuals of diverse ancestry in research on GLI.

## Materials and Methods

### Gene-lifestyle interaction (GLI) summary statistics

Summary statistics from four genome-wide interaction studies performed within the Cohorts for Heart and Aging Research in Genomic Epidemiology (CHARGE) consortium were gathered, covering blood pressure and lipid trait measures and cigarette smoking and alcohol consumption environmental exposures(6-10). The GLI models used additive allele effects and produced genome-wide joint tests of main effects and interaction effects. Sample sizes ranged from 175,000 to 602,000 individuals of multiple, self-identified ancestries (**Supplemental Table 1**).

We extracted individual variants with interaction p-value in any model less than 5×10^−5^ from each study for 5 subgroups, defined by the CHARGE Gene-Lifestyle Interaction Working Group: European ancestry (EA), African ancestry (AA), Hispanic ancestry (HA) and Asian Ancestry (ASA). Variants from trans-ancestry meta-analyses were also extracted for each project (**Supplemental Table 1**). Lipid trait measures included high-density lipoprotein cholesterol (HDL), low-density lipoprotein cholesterol (LDL), and triglycerides (TG). Four blood pressure traits were used: systolic blood pressure (SBP), diastolic BP (DBP), mean arterial pressure (MAP), and pulse pressure (PP). Environmental exposures were defined as: current drinking (yes/no), regular drinking (≥ 2 drinks per week/<2 drinks per week), drinking habits (≥ 8 glasses per week [heavy]/< 8 glasses per week [light]), current smoking (yes/no regular smoking in the past year), and ever smoked (yes/no 100 cigarettes smoked in lifetime).

### Epigenomic and transcriptomic data

Data describing the molecular signatures of the two environmental exposures were gathered from the literature, focusing on epigenomics and transcriptomics from population-based cohort studies (**Table 1**). For each of these sources, statistical significance as defined within the publication or resource was used for inclusion in this analysis. We used differential gene expression analysis (DExpr analysis) from studies of cigarette smoking(13, 17, 18) and alcohol consumption(15). For genes, locus boundaries were defined by the start of the first exon and the end of the final exon, regardless of transcript, extended by 500kb. Boundaries for DNA methylation (DNAm) sites were similarly defined by 500kb on either side. Differential methylation analyses (Dme analysis) yield DNAm sites whose methylation is associated with a trait or exposure. These sites can be further associated with gene expression, defining expression quantitative trait methylation sites (eQTM). DNAm sites and eQTM were gathered from studies for lipids(12), blood pressure(19), cigarette smoking(14), and alcohol consumption(15), with some also providing mQTL analyses results(15, 19). We also extracted significant expression quantitative trait loci (eQTL) from the Genetic Tissue-Expression (GTEx) project(16) portal, version 8.

### Locus prioritization based on accumulation of evidence

Our prioritization schema consisted of several steps. First, we identified overlapping loci from the GLI, Dme, eQTM and Dexpr analyses. Starting from a set of variants in the GLI loci, we found significantly associated DNAm sites within 500kb of the variants for the trait or exposure of interest. Next, we identified the genes for which both the GLI variants were significant eQTLs and the DNAm sites were significant eQTMs. Finally, we matched the phenotype or exposure results from the Dme results and selected genes with significant Dexpr effects.

## Data Availability

All data is publicly available from multiple, peer-reviewed publications. No individual-level measures were used in any analyses.

## Acknowledgments

This work was supported by the National Heart, Lung, and Blood Institute (NHLBI) of the National Institutes of Health (NIH) [R01 HL118305, R01 HL156991]; the Intramural Research Program of the National Human Genome Research Institute of the National Institutes of Health through the Center for Research on Genomics and Global Health (CRGGH) supported by the National Human Genome Research Institute, the National Institute of Diabetes and Digestive and Kidney Diseases, the Center for Information Technology, and the Office of the Director at the National Institutes of Health [Z01HG200362]; the American Heart Association [18CDA34110116 to PSdV]; and the National Institutes of Health [R01 DK117445, R01 MD012765, R21 HL140385 to NF].

## Terms

GLI: Gene-lifestyle interaction
LDL: low density lipoprotein
HDL: high density lipoprotein
TG: triglycerides
DBP: diastolic blood pressure
SBP: systolic blood pressure
MAP: mean arterial pressure
PP: pulse pressure
eQTL: expression quantitative trait loci
eQTM: expression quantitative trait methylation
mQTL: methylation quantitative trait loci
DNAm: DNA methylation
DMe: differential methylation
DExpr: differential gene expression

