## Supplemental Tables for "Multi-omics insights into the biological mechanisms underlying gene-by-lifestyle interactions with smoking and alcohol consumption detected by genome-wide trans-ancestry meta-analysis"

**Supplemental Table 1: CHARGE genome-wide interaction study overview.** Sample sizes represent combined Stage 1 (genome-wide) and Stage 2 (variants selected for follow-up).

| **Outcome** | | **Exposure** | **Sample Size** |
| --- | --- | --- | --- |
| Lipids | HDL | Current Drinking | 358K |
|  |  | Drinking Habits | 336K |
|  |  | Current Smoking | 386K |
|  |  | Ever Smoking | 385K |
|  | LDL | Current Drinking | 289K |
|  |  | Drinking Habits | 266K |
|  |  | Current Smoking | 313K |
|  |  | Ever Smoking | 311K |
|  | TG | Current Drinking | 326K |
|  |  | Drinking Habits | 313K |
|  |  | Current Smoking | 353K |
|  |  | Ever Smoking | 350K |
| Blood Pressure | SBP | Current Drinking | 548K |
|  |  | Drinking Habits | 176K |
|  |  | Current Smoking | 602K |
|  |  | Ever Smoking | 585K |
|  | DBP | Current Drinking | 548K |
|  |  | Drinking Habits | 176K |
|  |  | Current Smoking | 602K |
|  |  | Ever Smoking | 585K |
|  | MAP | Current Drinking | 548K |
|  |  | Drinking Habits | 175K |
|  |  | Current Smoking | 602K |
|  |  | Ever Smoking | 585K |
|  | PP | Current Drinking | 542K |
|  |  | Drinking Habits | 176K |
|  |  | Current Smoking | 601K |
|  |  | Ever Smoking | 584K |

**Supplemental Table 2.** **Prioritized loci with full eQTL information**

| Exposure | Gene Target | eQTL Tissue(s) | IntP < 5e-5 |
| --- | --- | --- | --- |
| *African Ancestry HDL* | | | |
| Curr Smk | POLK | Brain Cerebellum, Heart Left Ventricle, Nerve Tibial, Pancreas, Spleen, Thyroid, Brain Cerebellar Hemisphere | 5 |
| Curr Smk | *GCNT4^1^* | Artery Aorta, Testis | 4 |
| Curr Smk | *KLK8* | Skin Sun Exposed Lower leg | 4 |
| Curr Smk | *ANGPT1* | Thyroid | 3 |
| Curr Smk | *CARM1^2^* | Muscle Skeletal | 1 |
| Curr Smk | *LIPC* | Liver, Pancreas | 1 |
| Curr Smk | *YIPF2^2^* | Lung | 1 |
| Ev Smk | *PTPRZ1^1^*^,^*^2^* | Artery Aorta | 21 |
| Ev Smk | *DNAH7* | Adipose Subcutaneous, Adipose Visceral Omentum, Artery Tibial, Brain Caudate basal ganglia, Brain Cortex, Brain Nucleus accumbens basal ganglia, Brain Putamen basal ganglia, Breast Mammary Tissue, Cells Transformed fibroblasts, Esophagus Muscularis, Nerve Tibial, Ovary, Pancreas, Skin Not Sun Exposed Suprapubic, Skin Sun Exposed Lower leg, Stomach, Thyroid | 1 |
| *African Ancestry LDL* | | | |
| Ev Smk | *CRYGN* | Colon Sigmoid, Nerve Tibial | 1 |
| Ev Smk | *FRK* | Muscle Skeletal | 1 |
| Ev Smk | *WDR86* | Nerve Tibial | 1 |
| *African Ancestry TG* | | | |
| Curr Drnk | *BAHD1* | Colon Sigmoid, Esophagus Muscularis, Nerve Tibial, Pancreas, Stomach, Thyroid | 2 |
| Curr Drnk | *DNAJC17* | Esophagus Mucosa | 2 |
| Ev Smk | *TRIO* | Ovary | 1 |
| *African Ancestry DBP* | | | |
| Ev Smk | *FNTB* | Muscle Skeletal, Skin Sun Exposed Lower leg | 3 |
| Ev Smk | *RAB15* | Adipose Subcutaneous, Cells Transformed fibroblasts, Esophagus Muscularis, Nerve Tibial, Testis, Thyroid | 3 |
| *African Ancestry MAP* | | | |
| Curr Smk | *CRTAC1* | Adipose Visceral Omentum, Artery Tibial | 1 |
| Curr Smk | *SULT4A1* | Colon Sigmoid | 1 |
| Ev Smk | *FNTB* | Muscle Skeletal, Skin Sun Exposed Lower leg | 3 |
| Ev Smk | *RAB15* | Adipose Subcutaneous, Cells Transformed fibroblasts, Esophagus Muscularis, Nerve Tibial, Testis, Thyroid | 3 |
| Ev Smk | *KCNK13* | Brain Hypothalamus | 1 |
| *African Ancestry PP* | | | |
| Curr Drnk | *MPI* | Adipose Subcutaneous, Adipose Visceral Omentum, Artery Aorta, Artery Tibial, Brain Anterior cingulate cortex BA24, Brain Caudate basal ganglia, Brain Cerebellum, Brain Cortex, Brain Nucleus accumbens basal ganglia, Cells Transformed fibroblasts, Esophagus Mucosa, Esophagus Muscularis, Lung, Nerve Tibial, Skin Not Sun Exposed Suprapubic, Skin Sun Exposed Lower leg, Spleen, Testis, Thyroid, Whole Blood | 4 |
| Curr Drnk | *SCAMP2* | Esophagus Mucosa, Pituitary, Skin Sun Exposed Lower leg, Testis, Whole Blood | 4 |
| Curr Smk | *SYN2^1^* | Artery Aorta | 1 |
| Ev Smk | *JMJD4* | Nerve Tibial | 2 |
| *African Ancestry SBP* | | | |
| Curr Smk | *CRTAC1* | Adipose Visceral Omentum, Artery Tibial | 1 |
| Ev Smk | *FNTB* | Muscle Skeletal, Skin Sun Exposed Lower leg | 2 |
| Ev Smk | *RAB15* | Adipose Subcutaneous, Cells Transformed fibroblasts, Esophagus Muscularis, Nerve Tibial, Testis, Thyroid | 2 |
| Ev Smk | *JMJD4* | Nerve Tibial | 1 |
| *Asian Ancestry HDL* | | | |
| Ev Smk | *KCTD10^2^* | Artery Tibial, Skin Sun Exposed Lower leg | 1 |
| Ev Smk | *MYO1H^2^* | Lung | 1 |
| Ev Smk | *UBE3B^2^* | Colon Transverse, Skin Sun Exposed Lower leg | 1 |
| *Asian Ancestry LDL* | | | |
| Drnk habits | *CPNE2* | Cells Transformed fibroblasts | 3 |
| Ev Smk | *KANK2^2^* | Adipose Subcutaneous, Cells Transformed fibroblasts, Esophagus Muscularis, Lung, Nerve Tibial, Skin Sun Exposed Lower leg, Thyroid | 2 |
| *Asian Ancestry TG* | | | |
| Drnk habits | *LIPC^2^* | Thyroid | 1 |
| *Asian Ancestry MAP* | | | |
| Ev Smk | *ALDH2^1^*^,^*^2^* | Esophagus Mucosa, Skin Sun Exposed Lower leg, Lung, Nerve Tibial, Skin Not Sun Exposed Suprapubic, Thyroid | 2 |
| Ev Smk | *TMEM116^1^*^,^*^2^* | Adipose Visceral Omentum, Artery Tibial, Brain Frontal Cortex BA9, Esophagus Gastroesophageal Junction, Esophagus Mucosa, Esophagus Muscularis, Heart Atrial Appendage, Muscle Skeletal, Whole Blood-exp | 1 |
| *Asian Ancestry SBP* | | | |
| Curr Smk | *ALDH2^1^*^,^*^2^* | Esophagus Mucosa, Skin Sun Exposed Lower leg, Brain Cortex, Thyroid, Skin Not Sun Exposed Suprapubic | 3 |
| Ev Smk | *ALDH2^1^*^,^*^2^* | Esophagus Mucosa, Skin Sun Exposed Lower leg | 1 |
| *European Ancestry LDL* | | | |
| Drnk habits | *APOC1^2^* | Esophagus Mucosa | 1 |
| *European Ancestry TG* | | | |
| Drnk habits | *KRTCAP3^2^* | Adrenal Gland, Lung, Muscle Skeletal, Spleen, Thyroid, Whole Blood, Brain Caudate basal ganglia, Brain Cortex, Esophagus Mucosa, Skin Sun Exposed Lower leg | 2 |
| Drnk habits | *PPM1G^2^* | Esophagus Muscularis, Muscle Skeletal, Adipose Subcutaneous, Skin Sun Exposed Lower leg, Stomach, Thyroid | 2 |
| *Hispanic Ancestry PP* | | | |
| Curr Smk | *STIM1* | Lung, Thyroid, Whole Blood | 2 |
| *Trans Ancestry LDL* | | | |
| Ev Smk | *CRYGN* | Colon Sigmoid, Nerve Tibial | 1 |
| Ev Smk | *WDR86* | Nerve Tibial | 1 |
| *Trans Ancestry TG* | | | |
| Drnk habits | *KRTCAP3^2^* | Adrenal Gland, Lung, Muscle Skeletal, Spleen, Thyroid, Whole Blood | 1 |
| Drnk habits | *PPM1G^2^* | Esophagus Muscularis, Muscle Skeletal | 1 |

^1^Differential Expression by Exposure; ^2^Loci with 2df GLI association

**Supplemental Table 3: *GNCT4* × Current Smoking and HDL (African Ancestry)**

| **rsID** | **2df P-value** | **SNP Main Effect** | **Interaction P-value** | **Interaction Effect** | **eQTL Tissue** | **eQTL Effect** | **eQTL P-value** |
| --- | --- | --- | --- | --- | --- | --- | --- |
| rs11957913 | 0.0002049 | 0.003585 | 4.02E-05 | -0.0239 | Artery_Aorta | - | 7.95E-06 |
| rs11957913 | 0.0002049 | 0.003585 | 4.02E-05 | -0.0239 | Testis | + | 1.09E-13 |
| rs6886841 | 0.0003427 | 0.002521 | 3.45E-05 | -0.024 | Artery_Aorta | - | 2.96E-06 |
| rs6886841 | 0.0003427 | 0.002521 | 3.45E-05 | -0.024 | Testis | + | 2.45E-13 |
| rs3761743 | 0.0003735 | 0.002179 | 1.55E-05 | -0.0249 | Artery_Aorta | - | 3.52E-06 |
| rs3761743 | 0.0003735 | 0.002179 | 1.55E-05 | -0.0249 | Testis | + | 6.72E-14 |
| rs7737959 | 0.0006878 | 0.002938 | 3.65E-05 | -0.0241 | Artery_Aorta | - | 4.46E-06 |
| rs7737959 | 0.0006878 | 0.002938 | 3.65E-05 | -0.0241 | Testis | + | 1.01E-13 |
| rs7356637 | 0.0009189 | 0.003 | 4.88E-05 | -0.0237 | Artery_Aorta | - | 8.40E-06 |
| rs7356637 | 0.0009189 | 0.003 | 4.88E-05 | -0.0237 | Testis | + | 1.28E-13 |
| **Site name** | **P-value** | **Effect** | **Effect type** | | **Notes** | | |
| cg21158502 | 6.62E-06 | -0.006 | Smoking on methylation  (Difference in mean methylation) | | ARIC, FHS Offspring, KORA F4, GOLDN, LBC 1921, LBC 1936, NAS, Rotterdam, Inchianti, GTP, CHS European Ancestry (EA), CHS African Ancestry (AA), GENOA, EPIC Norfolk, EPIC, and MESA [Joehanes] | | |
| *GCNT4* | 9.88E-05 | 0.105 | Smoking on expression (Fold change) | | COPD Gene: self-identified Non- Hispanic Whites and African Americans between the ages of 45 and 80 years with a minimum of 10 pack-years life- time smoking history [Parker] | | |

**Supplemental Table 4: *PTPRZ1 ×* Ever Smoked and HDL (African Ancestry)**

| **rsID** | **2df P-value** | **SNP Main Effect** | **Interaction P-value** | **Interaction Effect** | **eQTL Tissue** | **eQTL Effect** | **eQTL P-value** |
| --- | --- | --- | --- | --- | --- | --- | --- |
| rs77810251 | 1.24E-09 | 0.05244 | 9.55E-07 | -0.0589 | Artery_Aorta | - | 1.47E-08 |
| rs6966138 | 5.53E-09 | 0.04933 | 2.42E-06 | -0.0555 | Artery_Aorta | - | 1.47E-08 |
| rs138601001 | 5.76E-09 | 0.04942 | 2.49E-06 | -0.0554 | Artery_Aorta | - | 1.57E-08 |
| rs3757551 | 8.63E-09 | 0.04895 | 3.02E-06 | -0.055 | Artery_Aorta | - | 1.47E-08 |
| rs6965735 | 9.41E-09 | -0.04875 | 3.30E-06 | 0.0548 | Artery_Aorta | + | 1.82E-08 |
| rs2109767 | 1.21E-08 | -0.04739 | 2.79E-06 | 0.054 | Artery_Aorta | + | 1.01E-08 |
| rs74755112 | 1.25E-08 | 0.04734 | 1.52E-06 | -0.0556 | Artery_Aorta | - | 1.47E-08 |
| rs75052642 | 1.48E-08 | 0.04815 | 4.52E-06 | -0.054 | Artery_Aorta | - | 1.47E-08 |
| rs740965 | 1.52E-08 | -0.0482 | 4.54E-06 | 0.054 | Artery_Aorta | + | 1.01E-08 |
| rs73229136 | 7.22E-07 | 0.02479 | 2.94E-06 | -0.0343 | Artery_Aorta | - | 2.59E-07 |
| rs3801377 | 1.49E-06 | -0.02648 | 4.44E-06 | 0.0363 | Artery_Aorta | + | 1.92E-07 |
| rs1916885 | 1.84E-06 | -0.02333 | 3.15E-06 | 0.034 | Artery_Aorta | + | 2.59E-07 |
| rs12669706 | 2.65E-06 | 0.02595 | 5.66E-06 | -0.0358 | Artery_Aorta | - | 1.92E-07 |
| rs56265902 | 2.69E-06 | 0.02611 | 5.31E-06 | -0.0359 | Artery_Aorta | - | 3.78E-07 |
| rs12672519 | 3.79E-06 | 0.02559 | 9.72E-06 | -0.0345 | Artery_Aorta | - | 1.63E-07 |
| rs12667535 | 5.35E-06 | 0.02473 | 6.61E-06 | -0.0353 | Artery_Aorta | - | 1.92E-07 |
| rs3801374 | 6.28E-06 | 0.02463 | 7.96E-06 | -0.035 | Artery_Aorta | - | 1.92E-07 |
| rs6971645 | 6.80E-06 | -0.02413 | 5.18E-06 | 0.0353 | Artery_Aorta | + | 1.92E-07 |
| rs3801373 | 7.01E-06 | 0.02447 | 8.03E-06 | -0.0349 | Artery_Aorta | - | 2.38E-07 |
| rs2024251 | 7.03E-06 | 0.02451 | 8.97E-06 | -0.0348 | Artery_Aorta | - | 1.92E-07 |
| rs9641678 | 1.24E-05 | -0.02355 | 9.09E-06 | 0.0346 | Artery_Aorta | + | 1.92E-07 |
| **Site name** | **Pvalue** | **Effect** | **Effect type** | | **Notes** | | |
| cg00826384 | 0.0011 | 1.90E-03 | Smoking on methylation (Difference in mean methylation) | | ARIC, FHS Offspring, KORA F4, GOLDN, LBC 1921, LBC 1936, NAS, Rotterdam, Inchianti, GTP, CHS European Ancestry (EA), CHS African Ancestry (AA), GENOA, EPIC Norfolk, EPIC, and MESA [Joehanes] | | |
| *PTPRZ1* | 0.041 | 0.9 | Smoking on expression (fold change) | | human neuroblastoma SH-SY5Y cells [Wang] | | |

**Supplemental Table 5: *SYN2 ×* Current Smoking on PP (African Ancestry)**

| **rsID** | **2df P-value** | **SNP Main Effect** | **Interaction P-value** | **Interaction Effect** | **eQTL Tissue** | **eQTL Effect** | **eQTL P-value** |
| --- | --- | --- | --- | --- | --- | --- | --- |
| rs4135300 | 8.72E-07 | 2.259 | 3.02E-07 | -9.8896 | Artery_Aorta | - | 2.19E-05 |
| **Site name** | **P-value** | **Effect** | **Effect type** | | **Notes** | | |
| cg10245988 | 9.00E-04 | 3.40E-03 | Smoking on methylation  (Difference in mean methylation) | | ARIC, FHS Offspring, KORA F4, GOLDN, LBC 1921, LBC 1936, NAS, Rotterdam, Inchianti, GTP, CHS European Ancestry (EA), CHS African Ancestry (AA), GENOA, EPIC Norfolk, EPIC, and MESA [Joehanes] | | |
| *SYN2* | 0.014 | 1.25 | Smoking on expression  (fold change) | | human neuroblastoma SH-SY5Y cells [Wang] | | |

**Supplemental Table 6: *ALDH2* × Ever Smoked and MAP (Asian Ancestry)**

| **rsID** | **2df P-value** | **SNP Main Effect** | **Interaction P-value** | **Interaction Effect** | **eQTL Tissue** | **eQTL Effect** | **eQTL P-value** |
| --- | --- | --- | --- | --- | --- | --- | --- |
| rs6490056 | 5.74E-11 | 0.1013 | 3.31E-05 | 0.5215 | Skin Sun Exposed Lower leg | - | 8.53E-08 |
| rs6490056 | 5.74E-11 | 0.1013 | 3.31E-05 | 0.5215 | Esophagus Mucosa | - | 2.42E-05 |
| rs10849962 | 2.62E-09 | 0.04002 | 4.69E-05 | 0.5046 | Lung | + | 5.76E-07 |
| rs10849962 | 2.62E-09 | 0.04002 | 4.69E-05 | 0.5046 | Nerve Tibial | + | 5.09E-07 |
| rs10849962 | 2.62E-09 | 0.04002 | 4.69E-05 | 0.5046 | Skin Not Sun Exposed Suprapubic | + | 3.48E-08 |
| rs10849962 | 2.62E-09 | 0.04002 | 4.69E-05 | 0.5046 | Esophagus Mucosa | + | 1.54E-10 |
| rs10849962 | 2.62E-09 | 0.04002 | 4.69E-05 | 0.5046 | Skin Sun Exposed Lower leg | + | 8.45E-12 |
| rs10849962 | 2.62E-09 | 0.04002 | 4.69E-05 | 0.5046 | Thyroid | + | 2.93E-06 |
| rs6490056 | 5.74E-11 | 0.1013 | 3.31E-05 | 0.5215 | Skin Sun Exposed Lower leg | - | 8.53E-08 |
| **Site name** | **P-value** | **Effect** | **Effect type** | | **Notes** | | |
| cg20884605 | 4.90E-07 | 4.30E-03 | Smoking on methylation (Difference in mean methylation) | | ARIC, FHS Offspring, KORA F4, GOLDN, LBC 1921, LBC 1936, NAS, Rotterdam, Inchianti, GTP, CHS European Ancestry (EA), CHS African Ancestry (AA), GENOA, EPIC Norfolk, EPIC, and MESA [Joehanes] | | |
| *ALDH2* | 9.68E-09 | 8.45E-02 | Smoking on expression (fold change) | | Framingham Heart Study (FHS), the Rotterdam Study (RS), the Cooperative Health Research in the Region of Augsburg (KORA F4) Study, the InCHIANTI Study, the Study of Health in Pomerania (SHIP-TREND), and the Estonian Biobank (EGCUT) [Huan] | | |

**Supplemental Table 7: *ALDH2 ×* Current Smoking and SBP (Asian Ancestry)**

| **rsID** | **2df P-value** | **SNP Main Effect** | **Interaction P-value** | **Interaction Effect** | **eQTL Tissue** | **eQTL Effect** | **eQTL P-value** |
| --- | --- | --- | --- | --- | --- | --- | --- |
| rs10774621 | 9.69E-13 | -0.3637 | 3.39E-05 | -0.8977 | Esophagus Mucosa | + | 2.04E-05 |
| rs10774621 | 9.69E-13 | -0.3637 | 3.39E-05 | -0.8977 | Skin Sun Exposed Lower leg | + | 2.06E-07 |
| rs10774621 | 9.69E-13 | -0.3637 | 3.39E-05 | -0.8977 | Brain Cortex | + | 4.45E-06 |
| rs10774621 | 9.69E-13 | -0.3637 | 3.39E-05 | -0.8977 | Thyroid | + | 1.04E-06 |
| rs73412370 | 1.23E-12 | 0.348 | 3.76E-05 | 0.8617 | Brain Cortex | - | 3.78E-06 |
| rs73412370 | 1.23E-12 | 0.348 | 3.76E-05 | 0.8617 | Esophagus Mucosa | - | 3.27E-05 |
| rs73412370 | 1.23E-12 | 0.348 | 3.76E-05 | 0.8617 | Skin Not Sun Exposed Suprapubic | - | 8.35E-06 |
| rs73412370 | 1.23E-12 | 0.348 | 3.76E-05 | 0.8617 | Skin Sun Exposed Lower leg | - | 2.36E-07 |
| rs73412370 | 1.23E-12 | 0.348 | 3.76E-05 | 0.8617 | Thyroid | - | 2.82E-08 |
| rs6490056 | 1.99E-10 | 0.2777 | 2.75E-05 | 0.9462 | Esophagus Mucosa | - | 2.42E-05 |
| rs6490056 | 1.99E-10 | 0.2777 | 2.75E-05 | 0.9462 | Skin Sun Exposed Lower leg | - | 8.53E-08 |
| rs10774621 | 9.69E-13 | -0.3637 | 3.39E-05 | -0.8977 | Esophagus Mucosa | + | 2.04E-05 |
| **Site name** | **P-value** | **Effect** | **Effect type** | | **Notes** | | |
| cg20884605 | 4.90E-07 | 4.30E-03 | Smoking on methylation (Difference in mean methylation) | | ARIC, FHS Offspring, KORA F4, GOLDN, LBC 1921, LBC 1936, NAS, Rotterdam, Inchianti, GTP, CHS European Ancestry (EA), CHS African Ancestry (AA), GENOA, EPIC Norfolk, EPIC, and MESA [Joehanes] | | |
| *ALDH2* | 9.68E-09 | 8.45E-02 | Smoking on expression  (fold change) | | Framingham Heart Study (FHS), the Rotterdam Study (RS), the Cooperative Health Research in the Region of Augsburg (KORA F4) Study, the InCHIANTI Study, the Study of Health in Pomerania (SHIP-TREND), and the Estonian Biobank (EGCUT) [Huan] | | |

**Supplemental Table 8: *ALDH2 ×* Ever Smoked and SBP (Asian Ancestry)**

| **rsID** | **2df P-value** | **SNP Main Effect** | **Interaction P-value** | **Interaction Effect** | **eQTL Tissue** | **eQTL Effect** | **eQTL P-value** |
| --- | --- | --- | --- | --- | --- | --- | --- |
| rs6490056 | 3.42E-10 | 0.1131 | 3.78E-05 | 0.7353 | Esophagus Mucosa | - | 2.42E-05 |
| rs6490056 | 3.42E-10 | 0.1131 | 3.78E-05 | 0.7353 | Skin Sun Exposed Lower leg | - | 8.53E-08 |
| **Site name** | **P-value** | **Effect** | **Effect type** | | **Notes** | | |
| cg20884605 | 4.90E-07 | 4.30E-03 | Smoking on methylation (Difference in mean methylation) | | ARIC, FHS Offspring, KORA F4, GOLDN, LBC 1921, LBC 1936, NAS, Rotterdam, Inchianti, GTP, CHS European Ancestry (EA), CHS African Ancestry (AA), GENOA, EPIC Norfolk, EPIC, and MESA [Joehanes] | | |
| *ALDH2* | 9.68E-09 | 8.45E-02 | Smoking on expression (fold change) | | Framingham Heart Study (FHS), the Rotterdam Study (RS), the Cooperative Health Research in the Region of Augsburg (KORA F4) Study, the InCHIANTI Study, the Study of Health in Pomerania (SHIP-TREND), and the Estonian Biobank (EGCUT) [Huan] | | |

**Supplemental Table 9: *TMEM116 ×* Ever Smoked and MAP (Asian Ancestry)**

| **rsID** | **2df Pvalue** | **SNP main Effect** | **Interaction Pvalue** | **Interaction Effect** | **eQTL Tissue** | **eQTL Effect** | **eQTL Pvalue** |
| --- | --- | --- | --- | --- | --- | --- | --- |
| rs6490056 | 5.74E-11 | 0.1013 | 3.31E-05 | 0.5215 | Heart Atrial Appendage | + | 8.59E-06 |
| rs6490056 | 5.74E-11 | 0.1013 | 3.31E-05 | 0.5215 | Adipose Visceral Omentum | + | 5.88E-06 |
| rs10849962 | 2.62E-09 | 0.04002 | 4.69E-05 | 0.5046 | Esophagus Gastroesophageal Junction | - | 2.83E-06 |
| rs10849962 | 2.62E-09 | 0.04002 | 4.69E-05 | 0.5046 | Adipose Visceral Omentum | - | 7.58E-07 |
| rs10849962 | 2.62E-09 | 0.04002 | 4.69E-05 | 0.5046 | Artery Tibial | - | 2.50E-05 |
| rs10849962 | 2.62E-09 | 0.04002 | 4.69E-05 | 0.5046 | Brain Frontal Cortex BA9 | - | 3.62E-05 |
| rs10849962 | 2.62E-09 | 0.04002 | 4.69E-05 | 0.5046 | Heart Atrial Appendage | - | 2.03E-07 |
| rs10849962 | 2.62E-09 | 0.04002 | 4.69E-05 | 0.5046 | Esophagus Mucosa | - | 6.11E-05 |
| rs10849962 | 2.62E-09 | 0.04002 | 4.69E-05 | 0.5046 | Esophagus Muscularis | - | 6.87E-06 |
| rs10849962 | 2.62E-09 | 0.04002 | 4.69E-05 | 0.5046 | Muscle Skeletal | - | 1.25E-06 |
| rs10849962 | 2.62E-09 | 0.04002 | 4.69E-05 | 0.5046 | Whole Blood | - | 2.98E-05 |
| **Site name** | **P-value** | **Effect** | **Effect type** | | **Notes** | | |
| cg08528204 | 2.07E-08 | 0.03 | Smoking on methylation (Difference in mean methylation) | | Family and Community Health Study (FACHS) (African American females from Iowa and Georgia, n=111) [Huan] | | |
| *TMEM116* | 8.56E-04 | 3.76E-02 | Smoking on expression (fold change) | | Framingham Heart Study (FHS), the Rotterdam Study (RS), the Cooperative Health Research in the Region of Augsburg (KORA F4) Study, the InCHIANTI Study, the Study of Health in Pomerania (SHIP-TREND), and the Estonian Biobank (EGCUT)  [Huan] | | |
