## Supplemental Figures for "Multi-omics insights into the biological mechanisms underlying gene-by-lifestyle interactions with smoking and alcohol consumption detected by genome-wide trans-ancestry meta-analysis"

**Supplemental Figure 1.** Manhattan plots of GLI association results with lipids (top) and blood pressure (bottom) in (A) trans-ancestry meta analyses, (B) African Ancestry subgroup, (C) Asian Ancestry subgroup, (D) European Ancestry subgroup, and (E) Hispanic Ancestry subgroup analyses.

A.

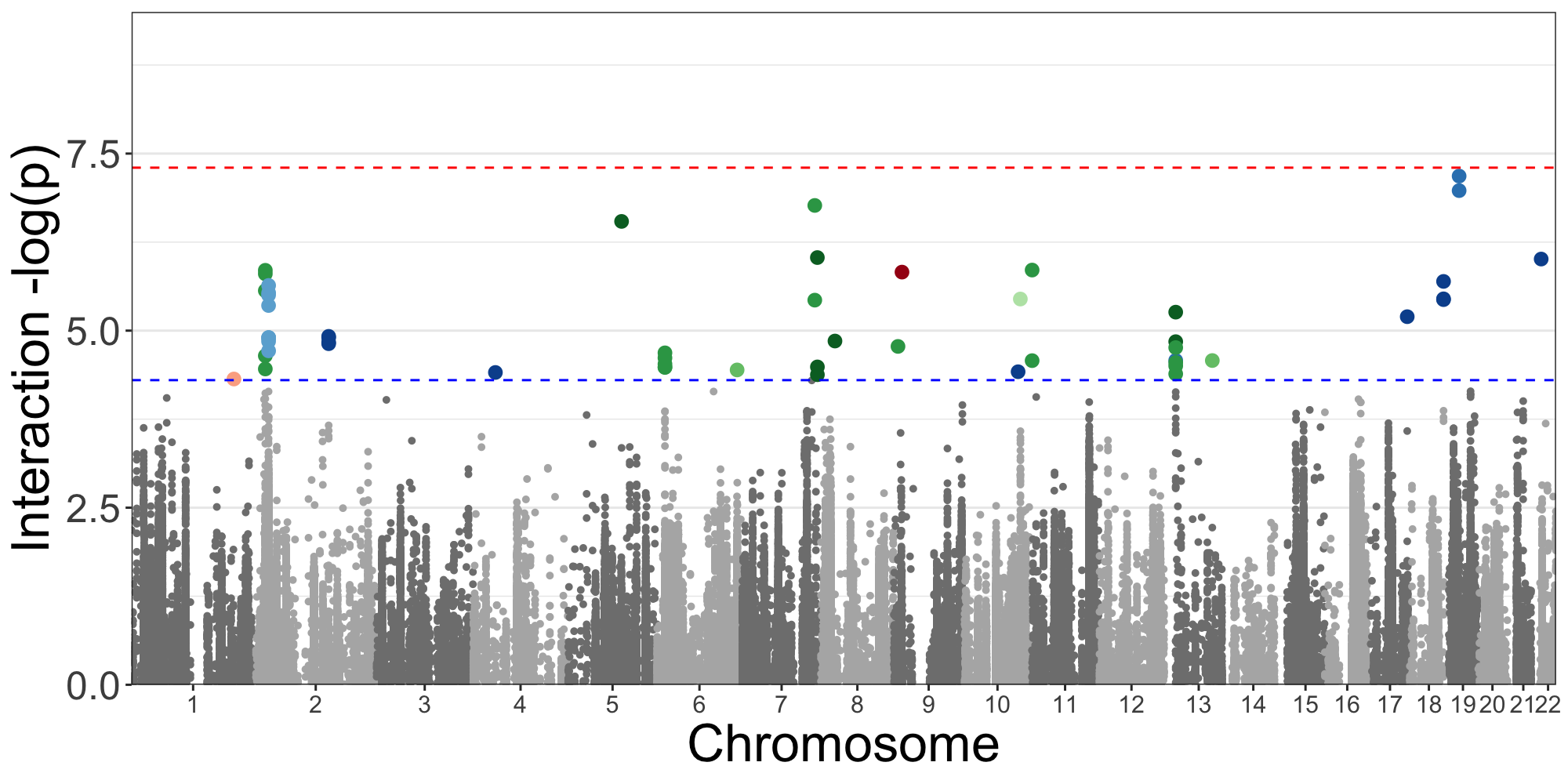

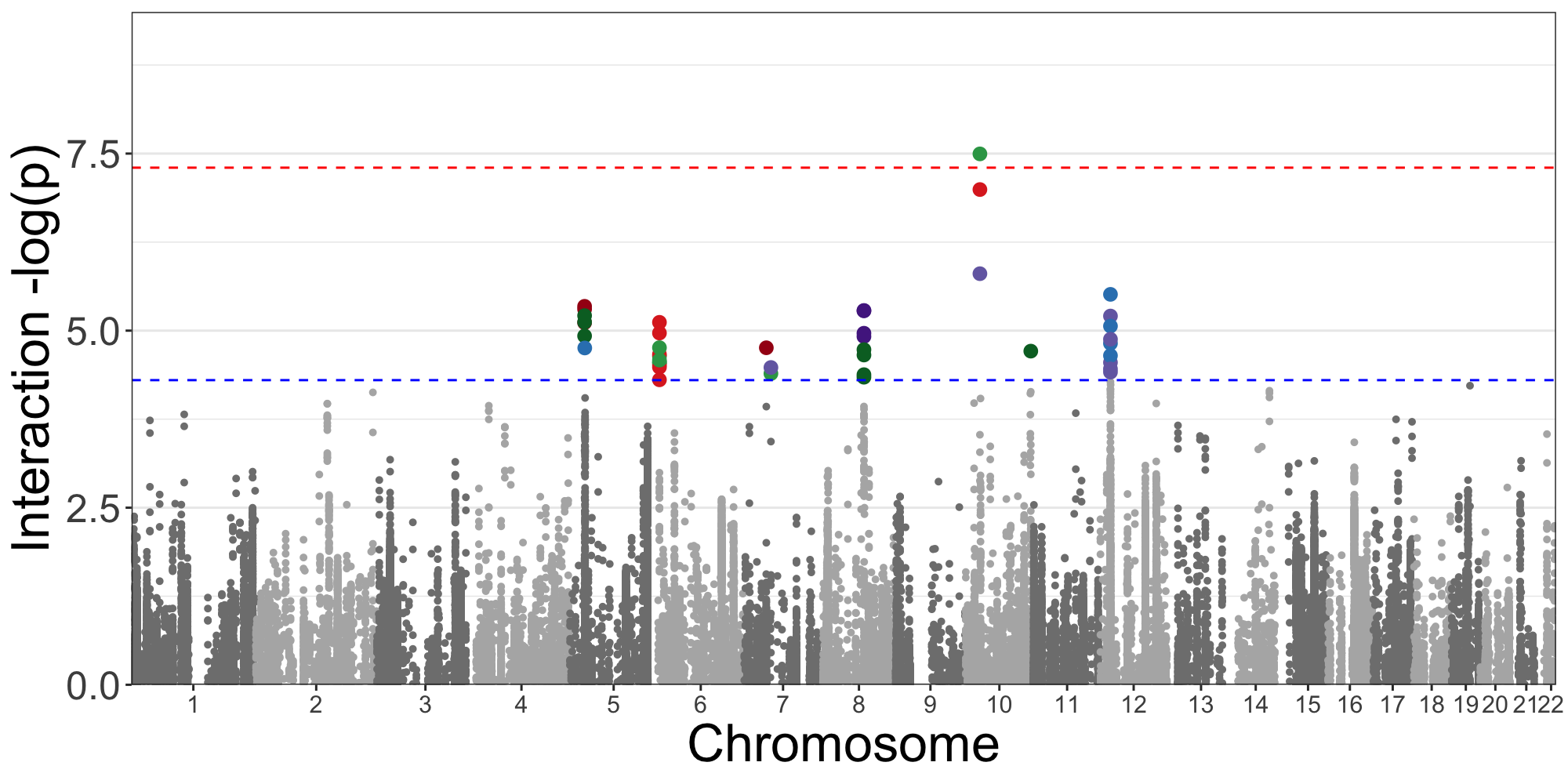

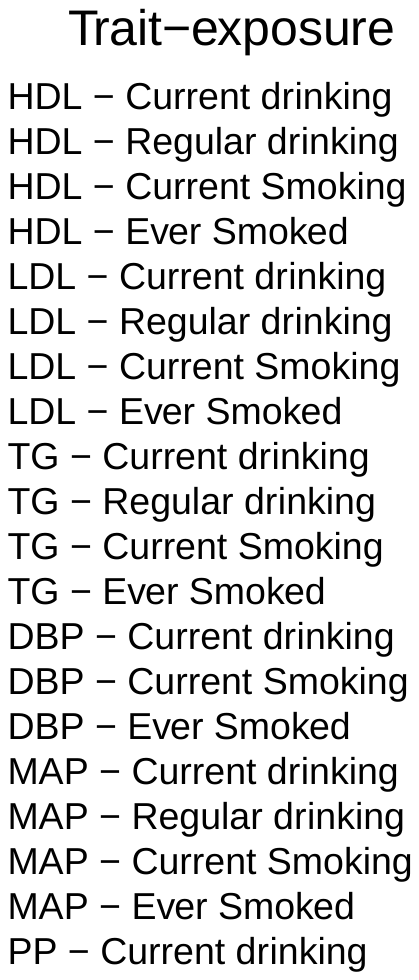

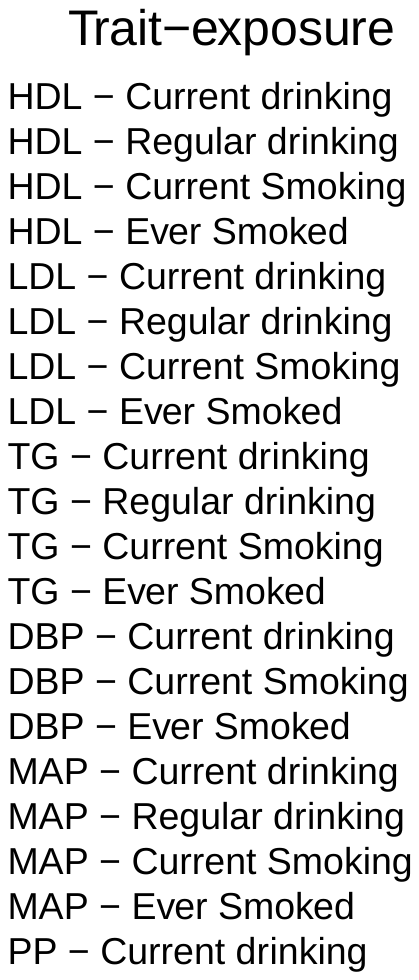

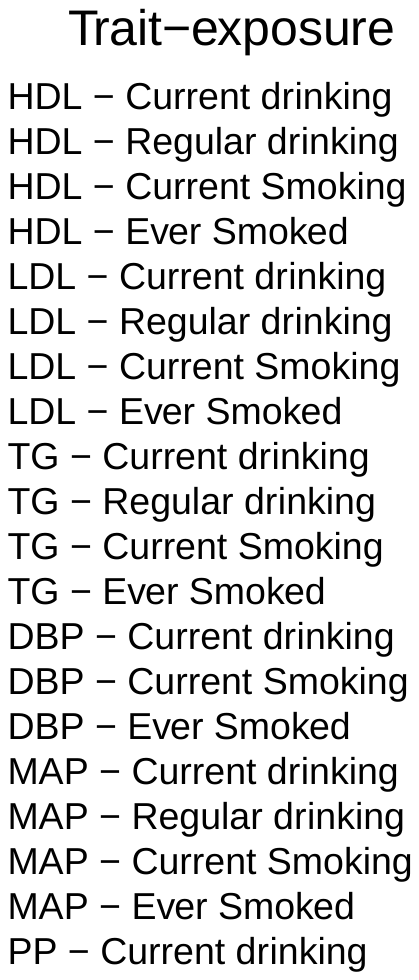

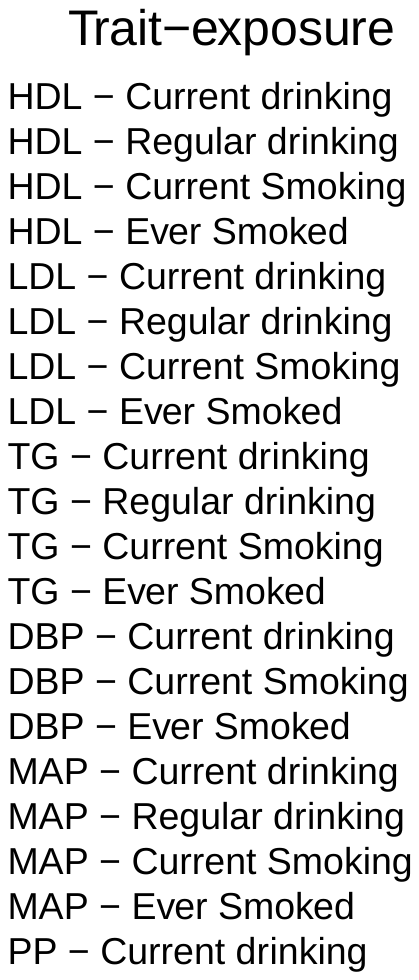

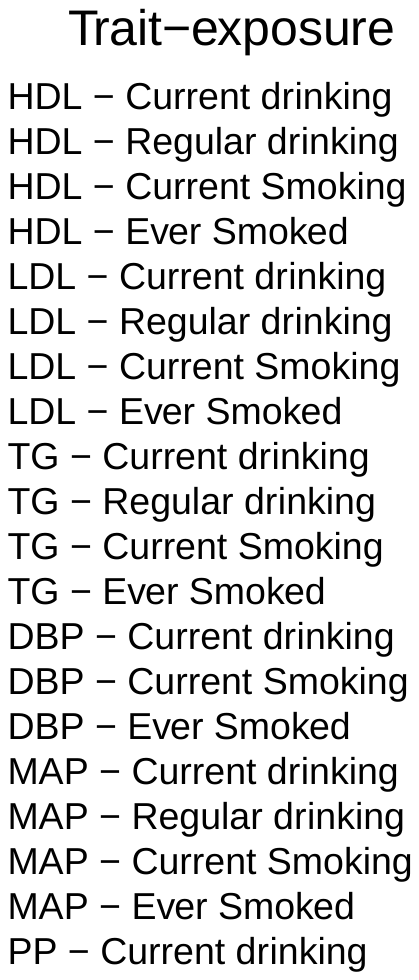

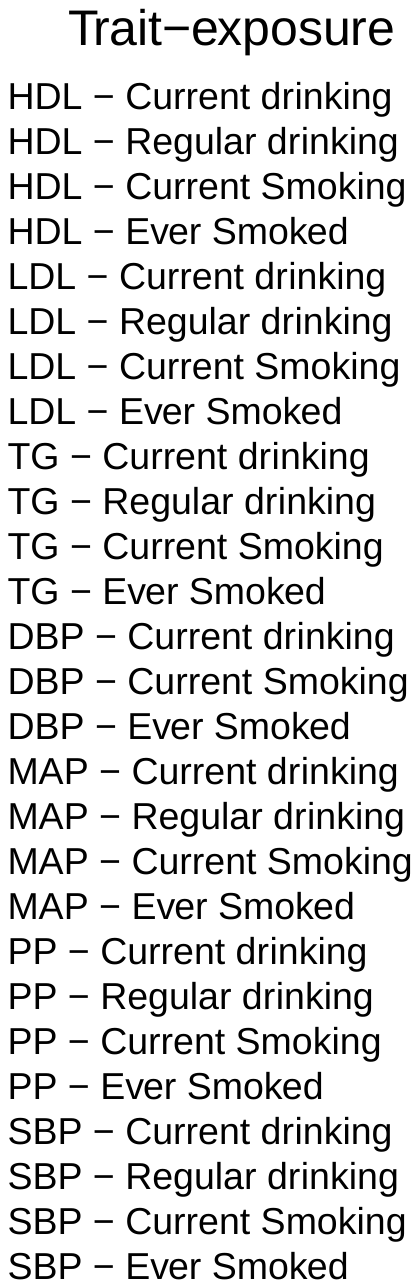

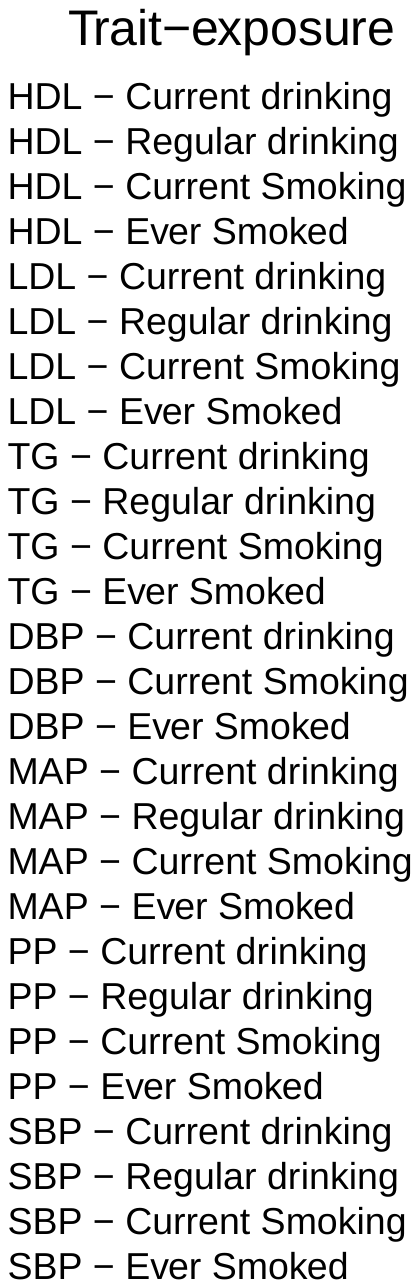

B.

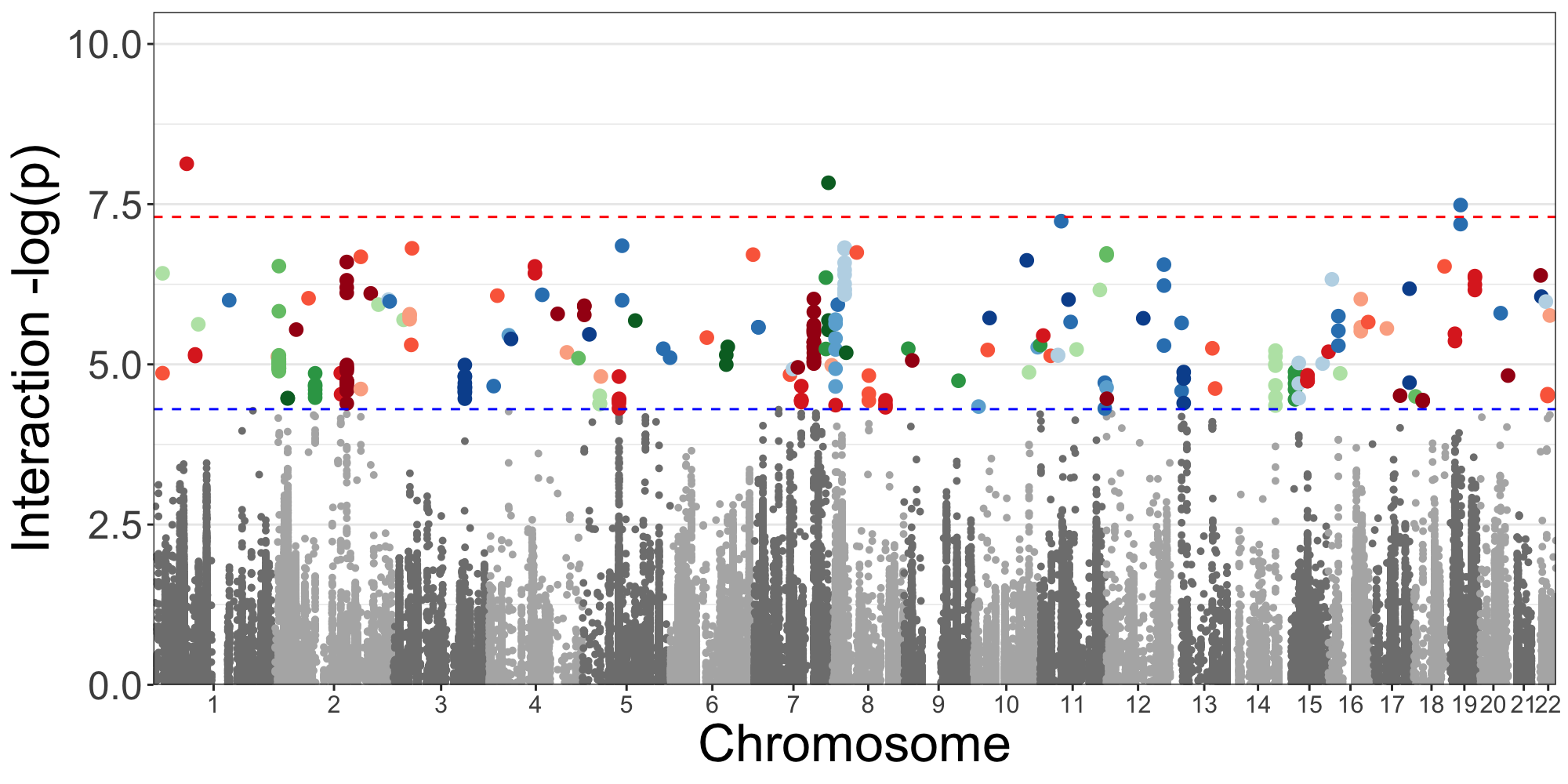

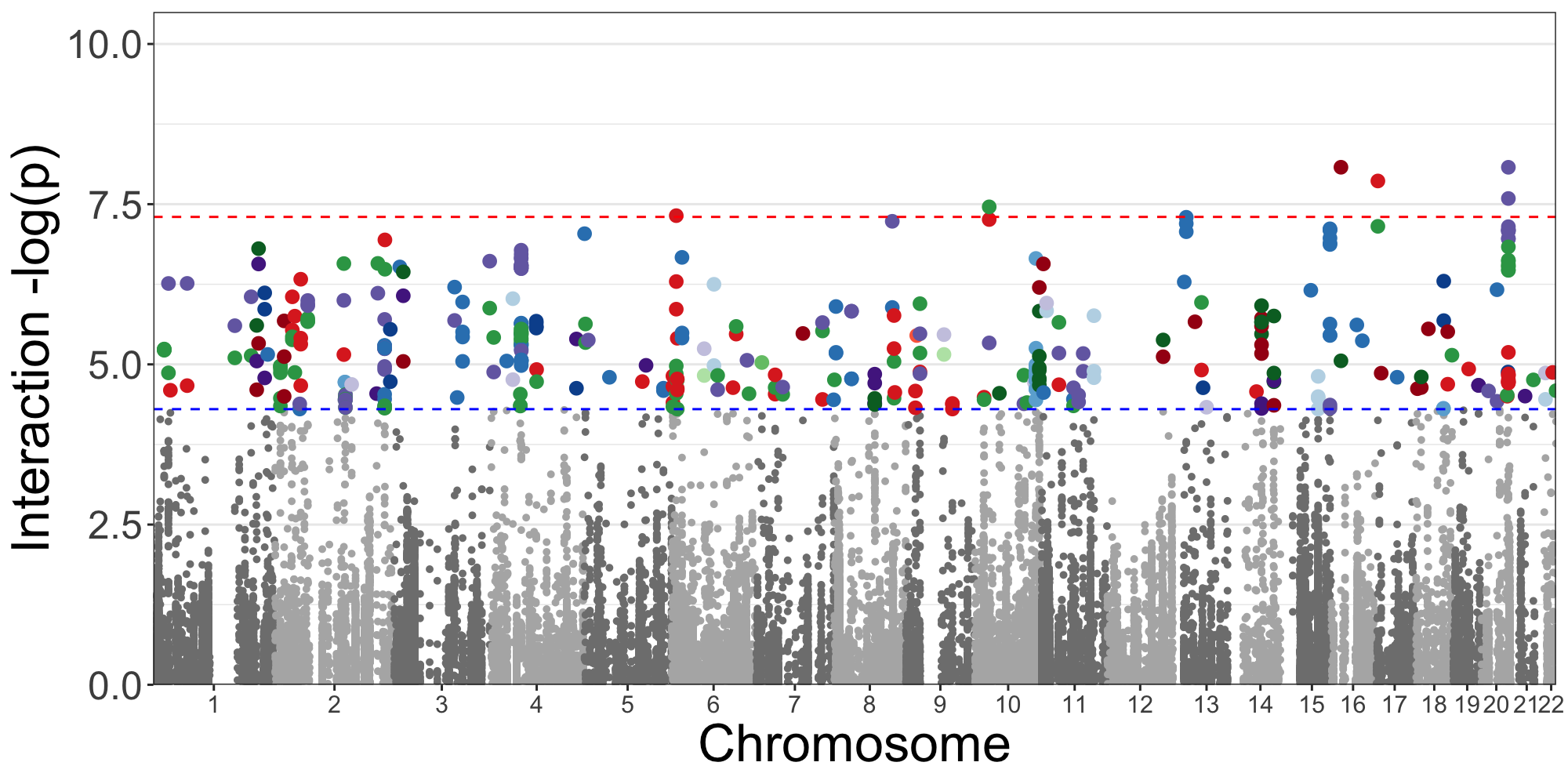

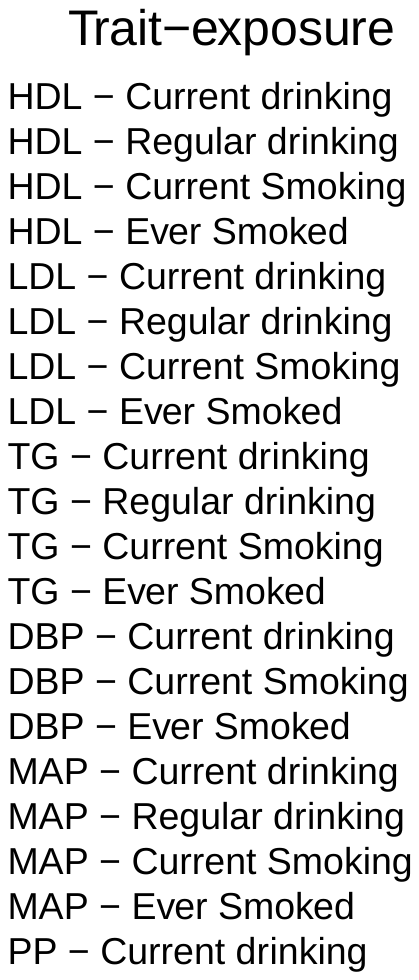

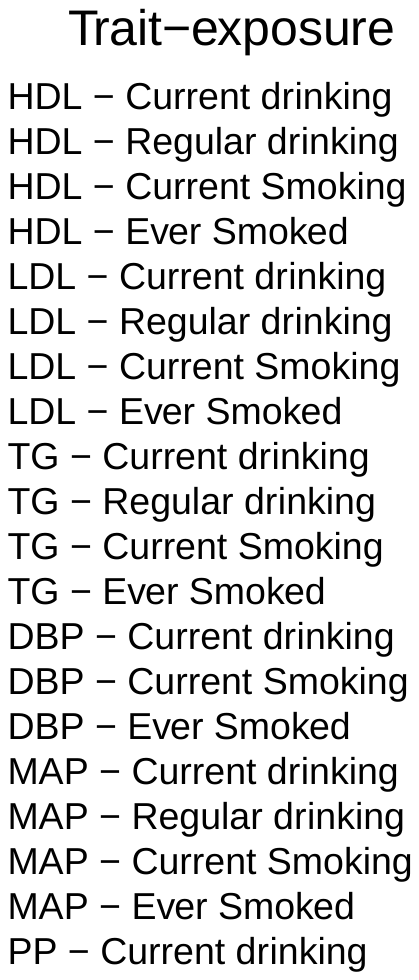

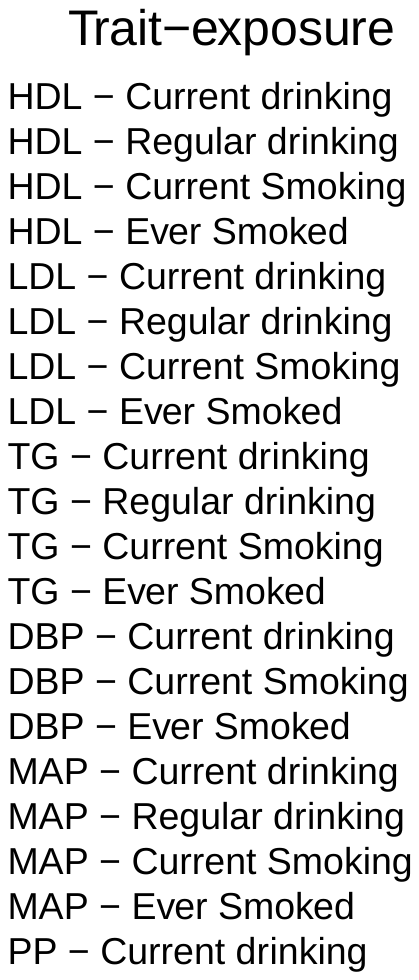

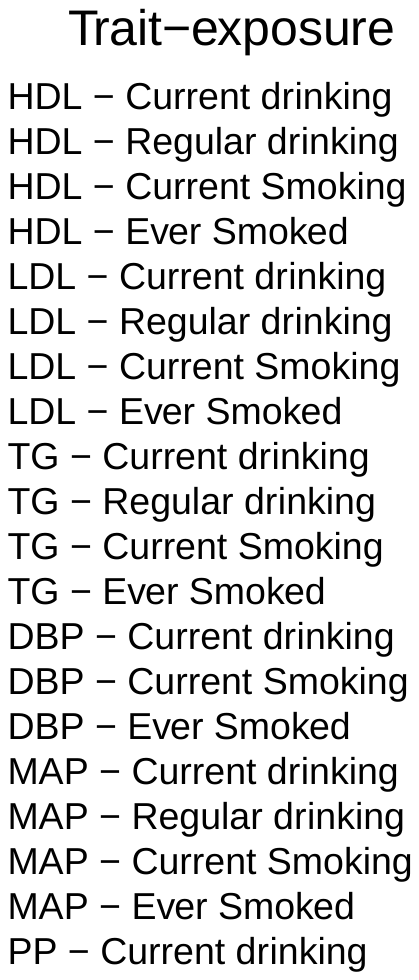

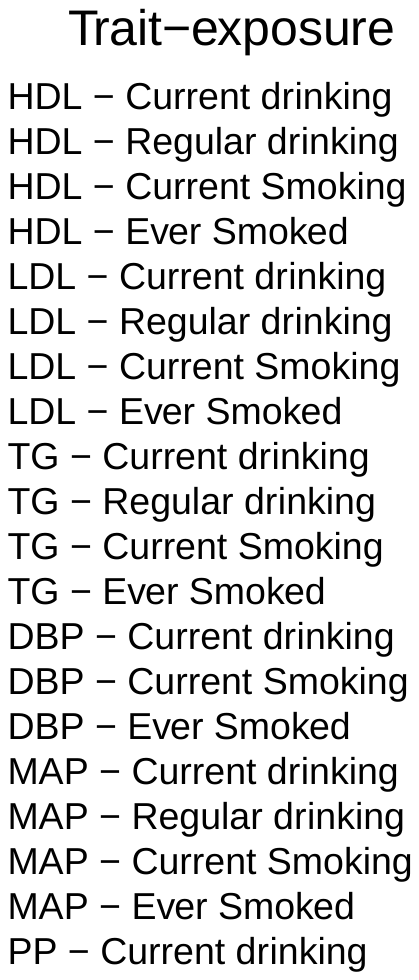

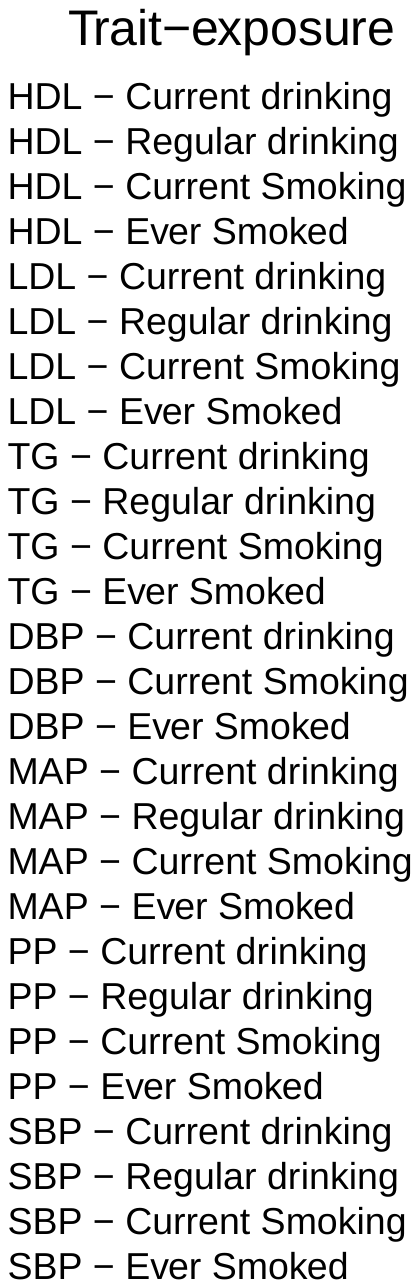

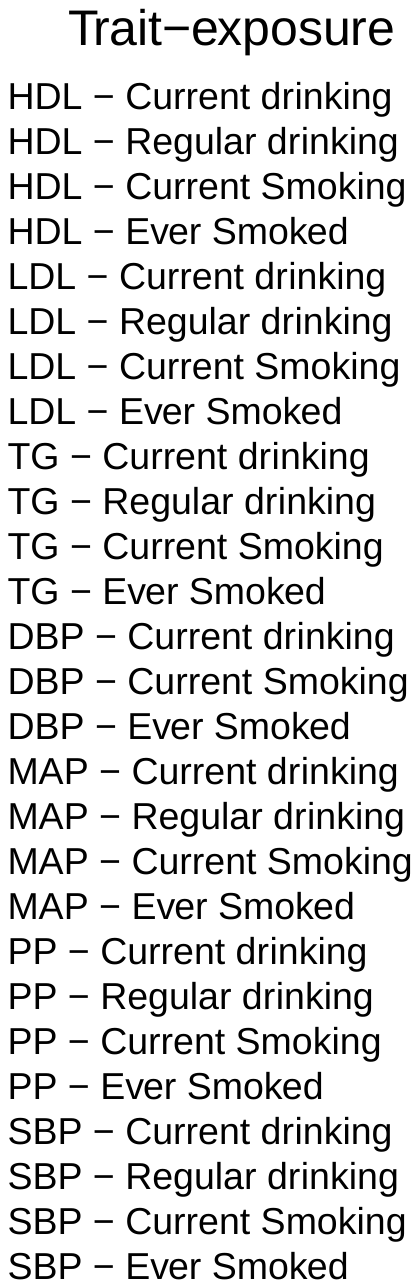

C.

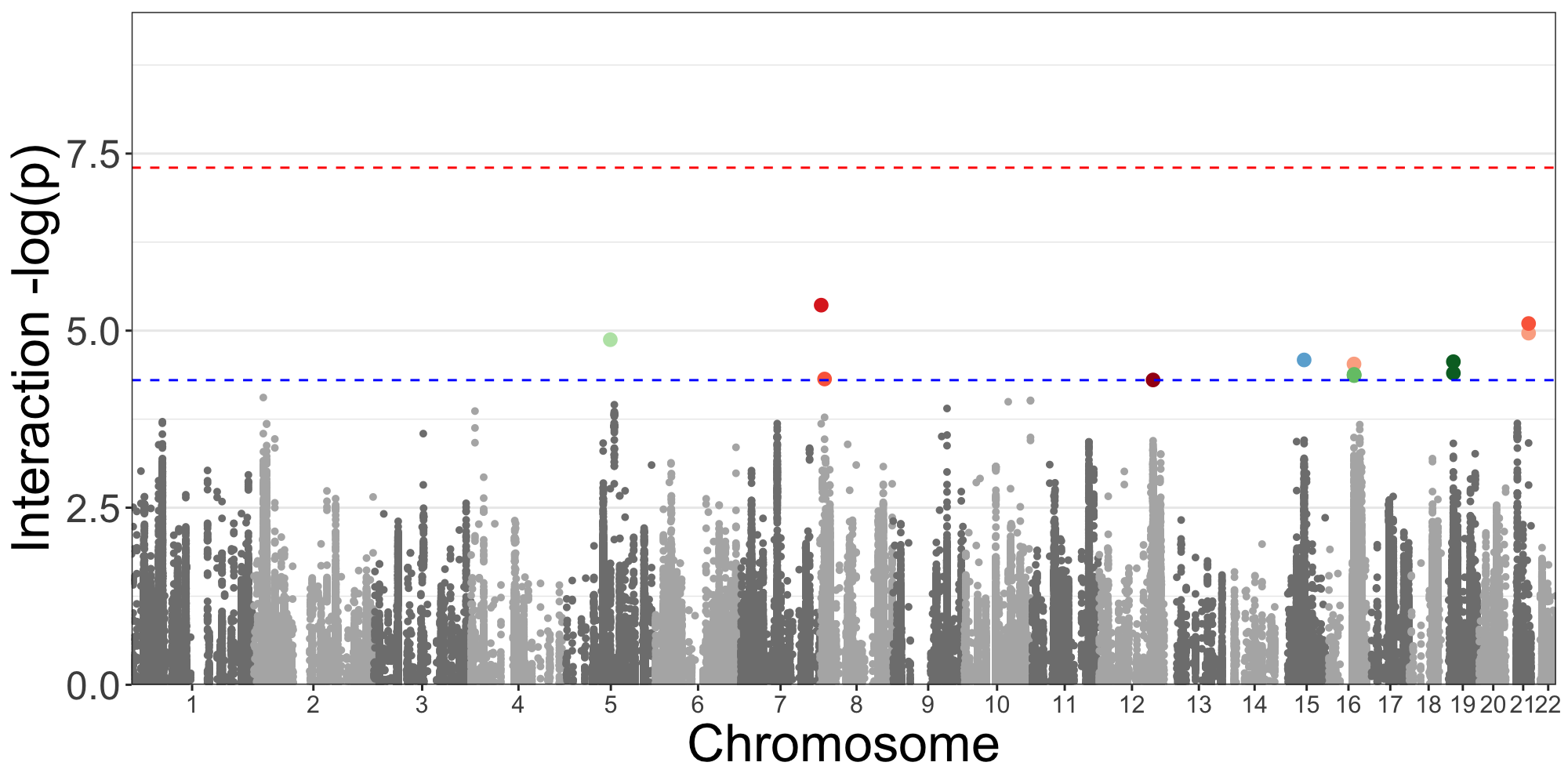

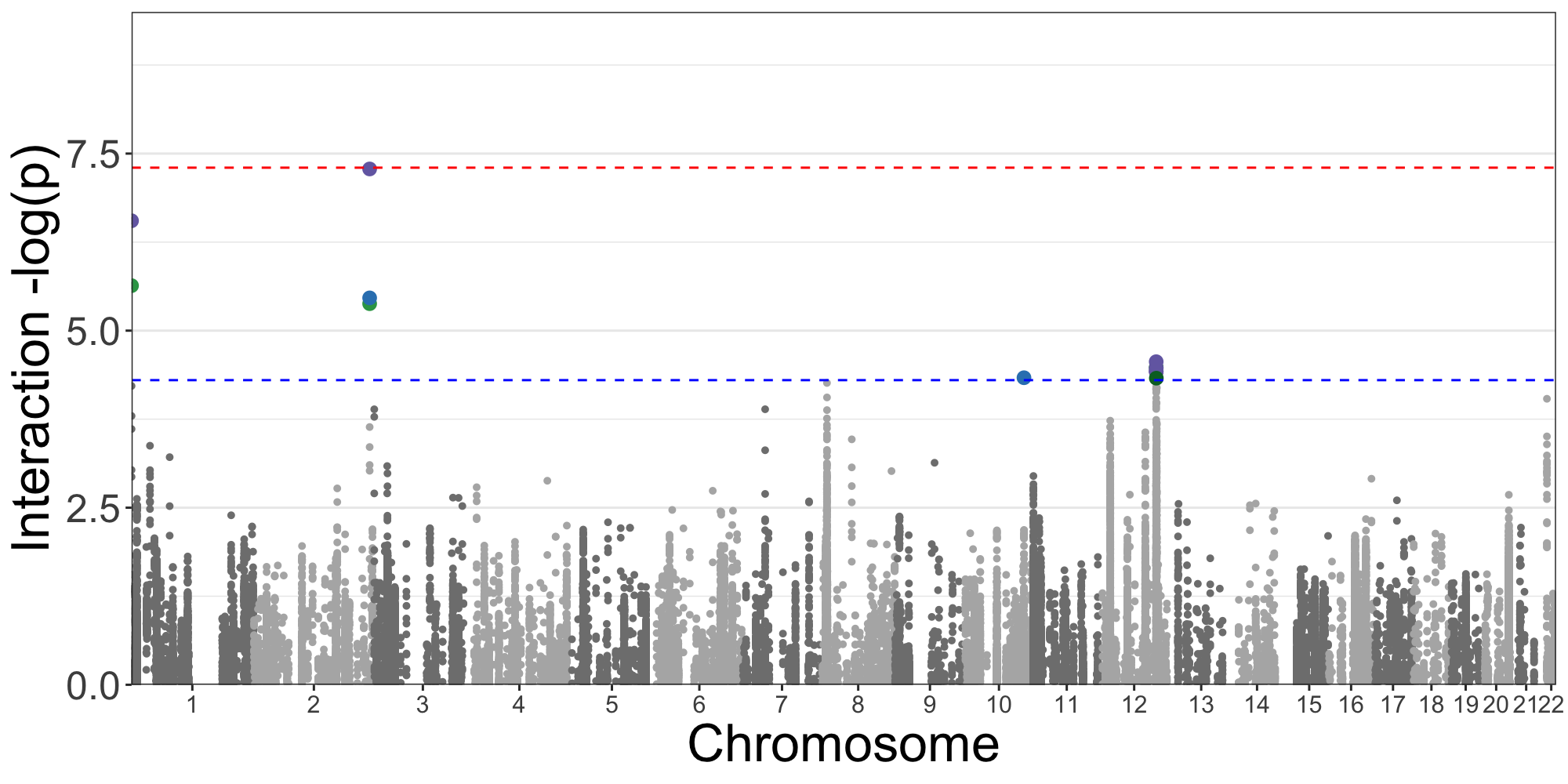

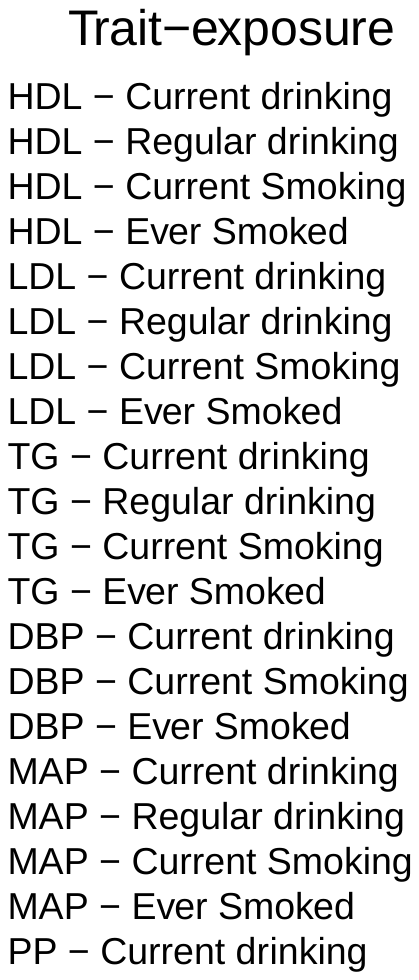

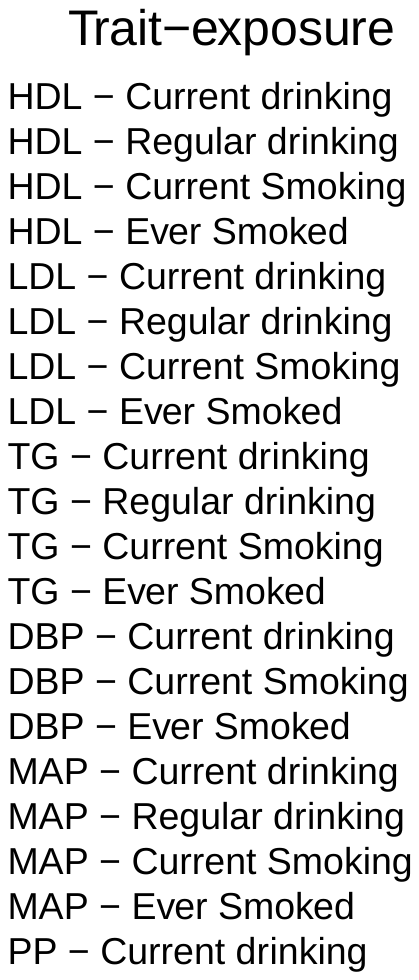

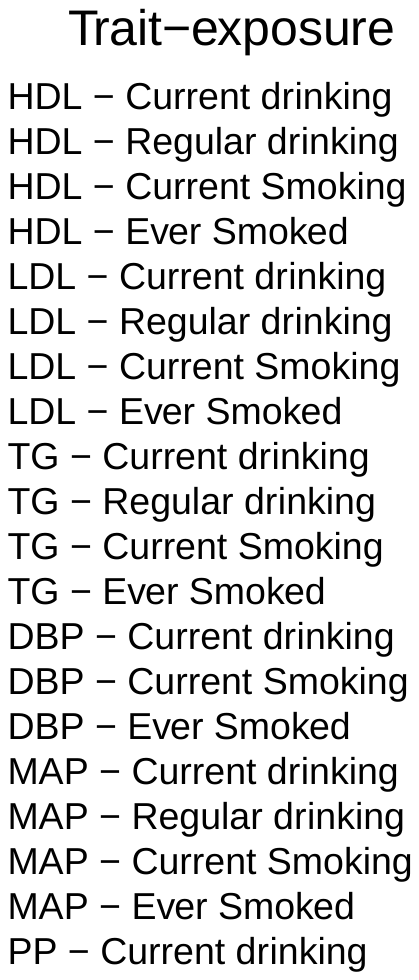

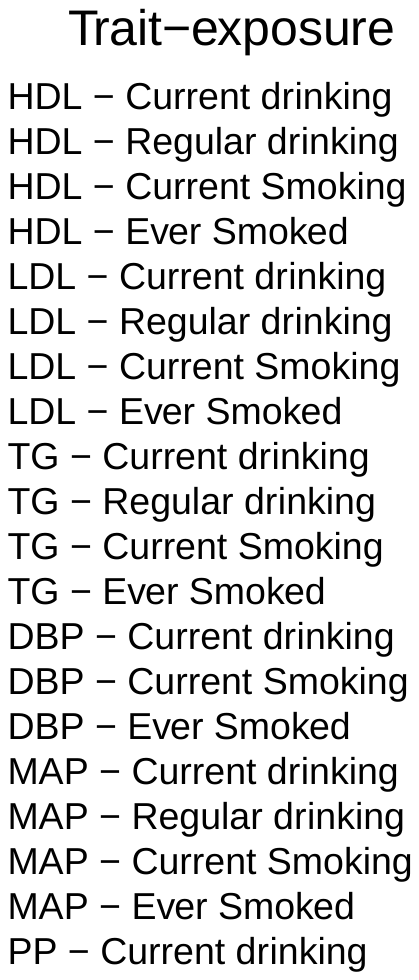

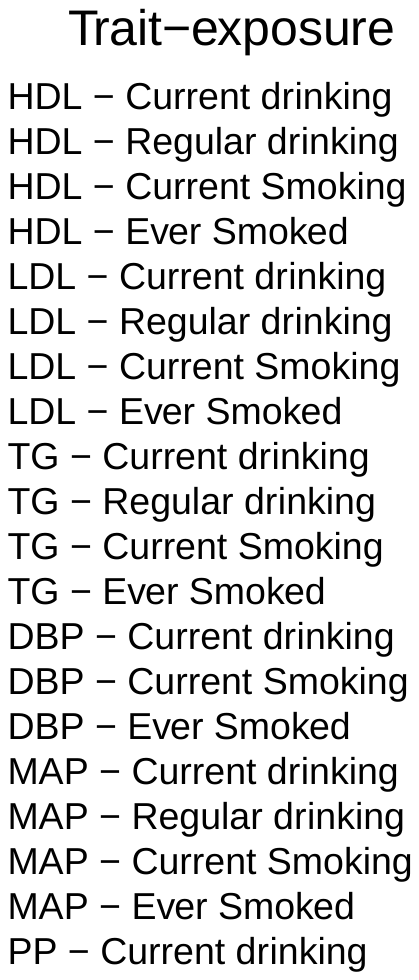

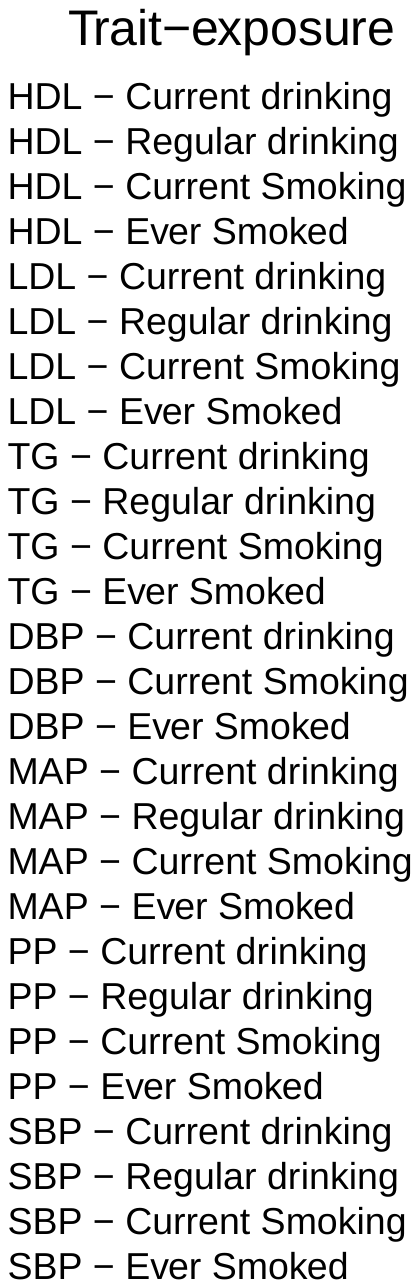

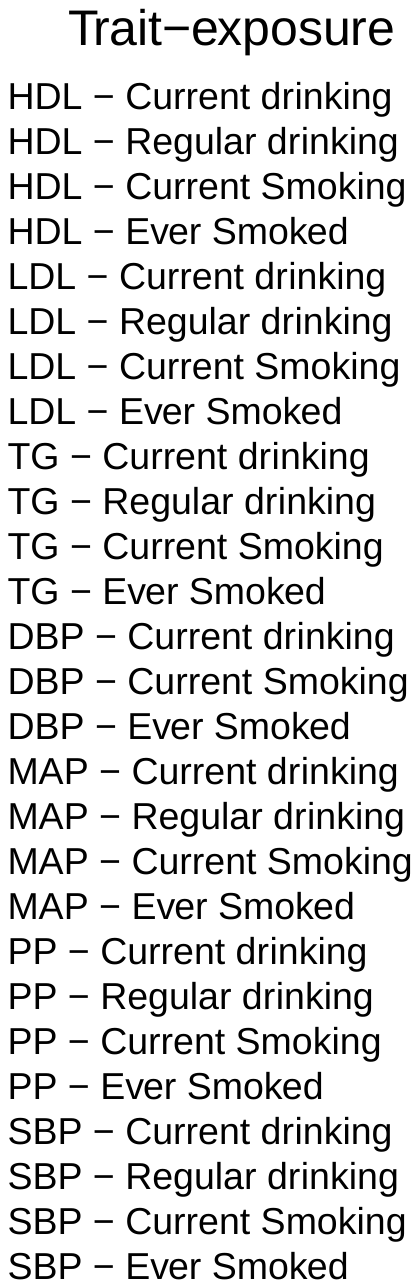

D.

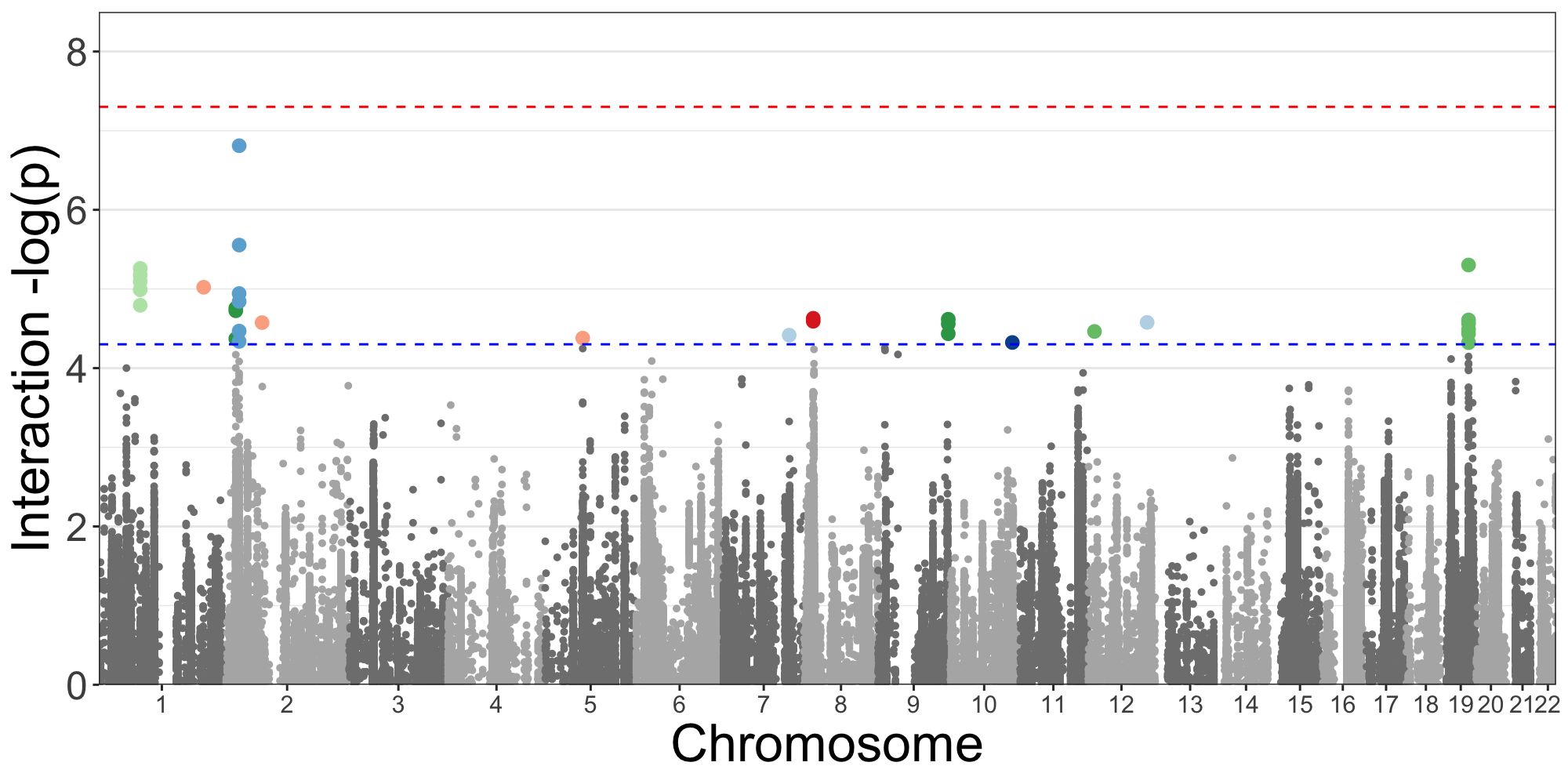

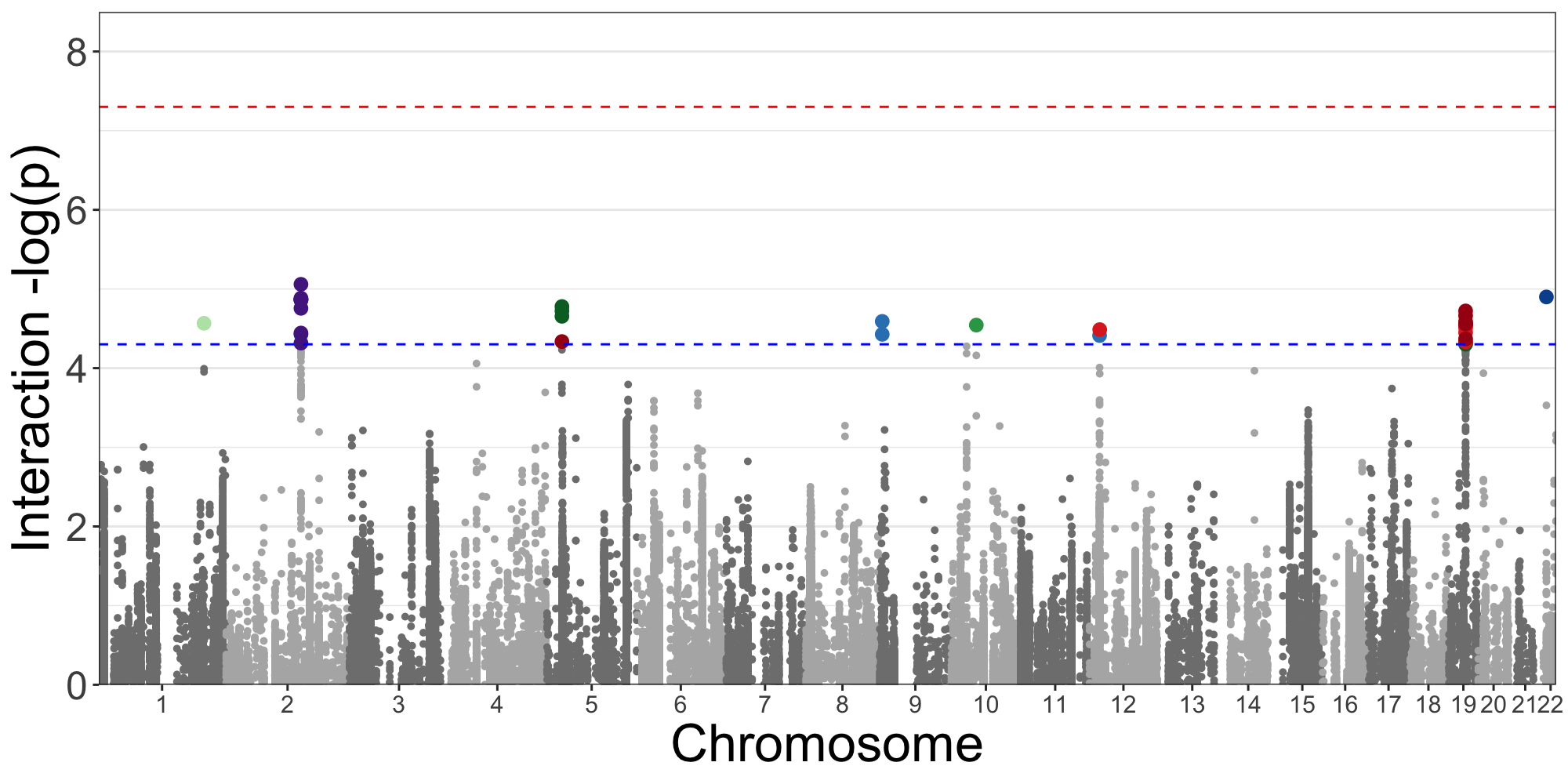

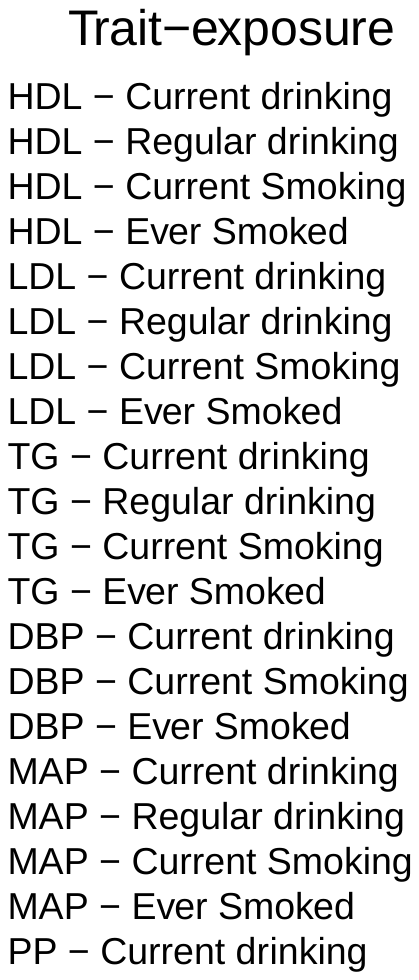

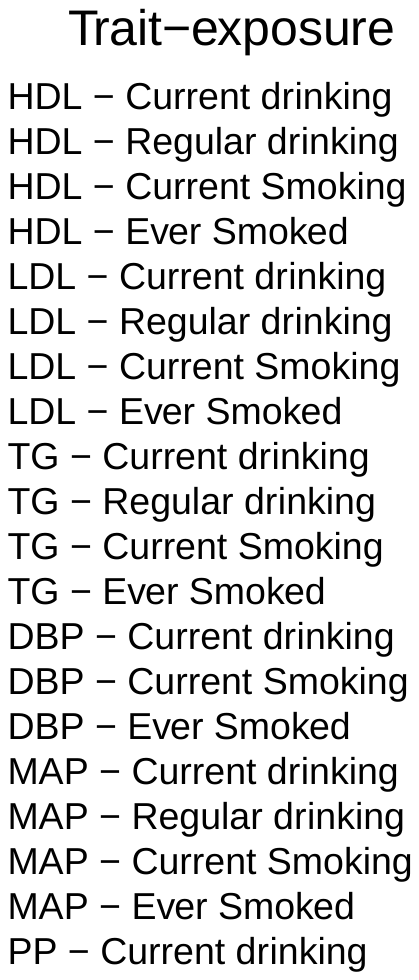

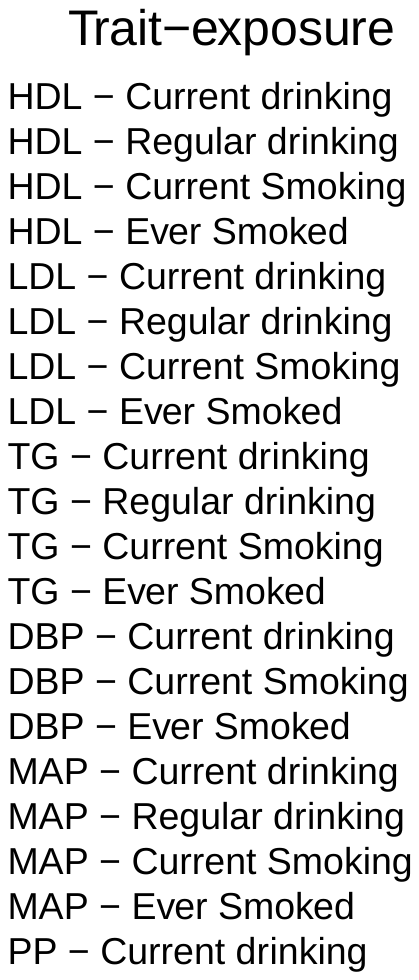

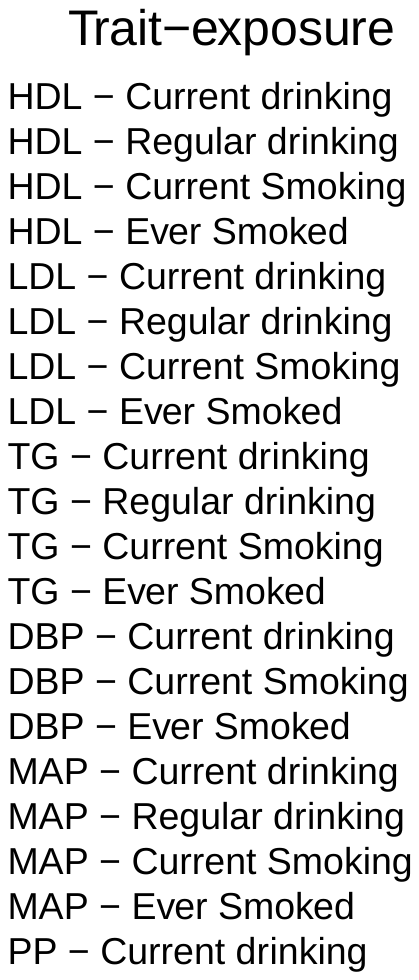

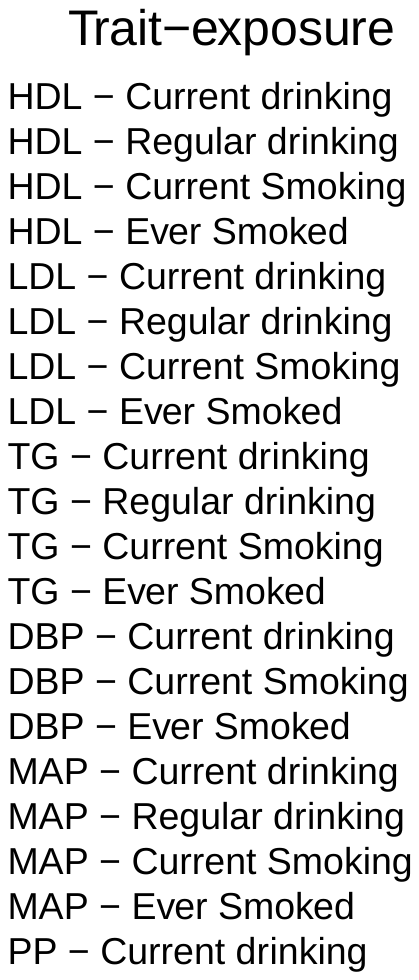

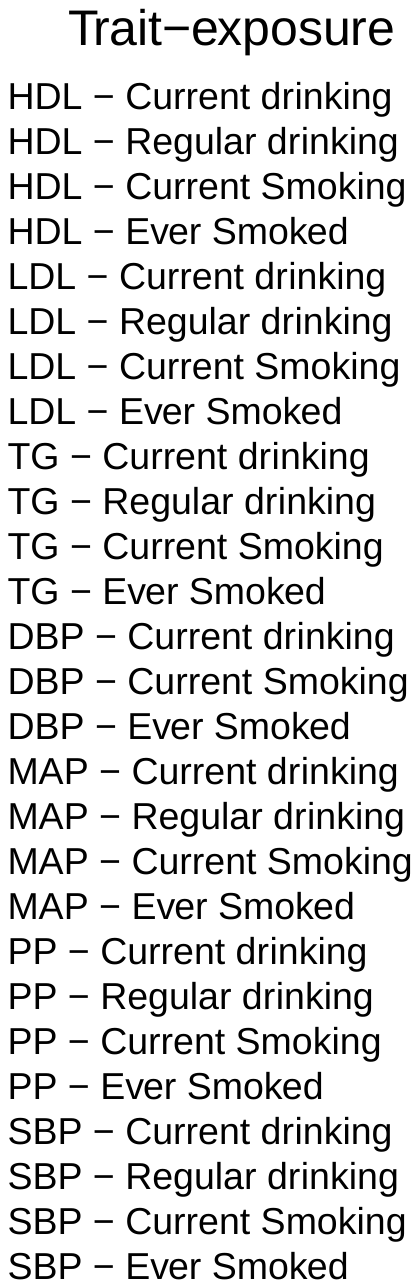

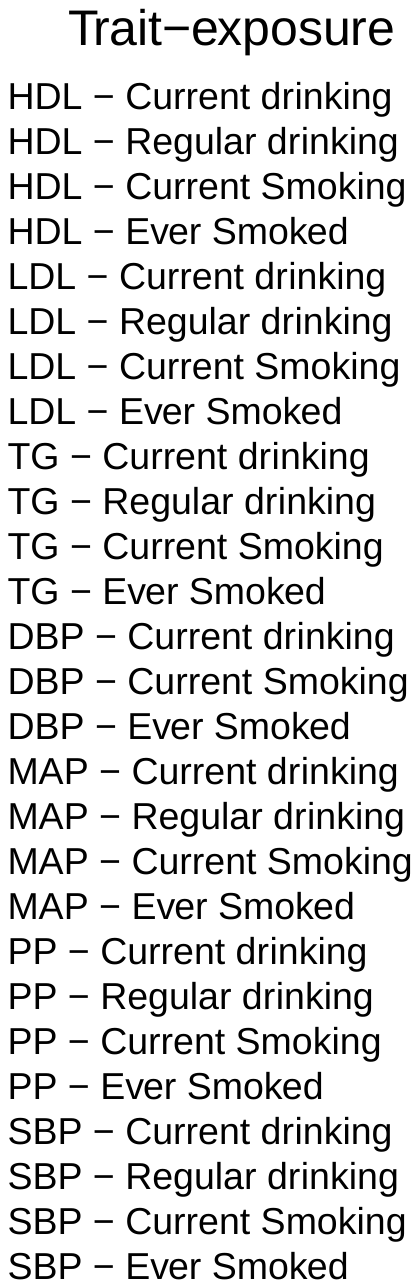

E.

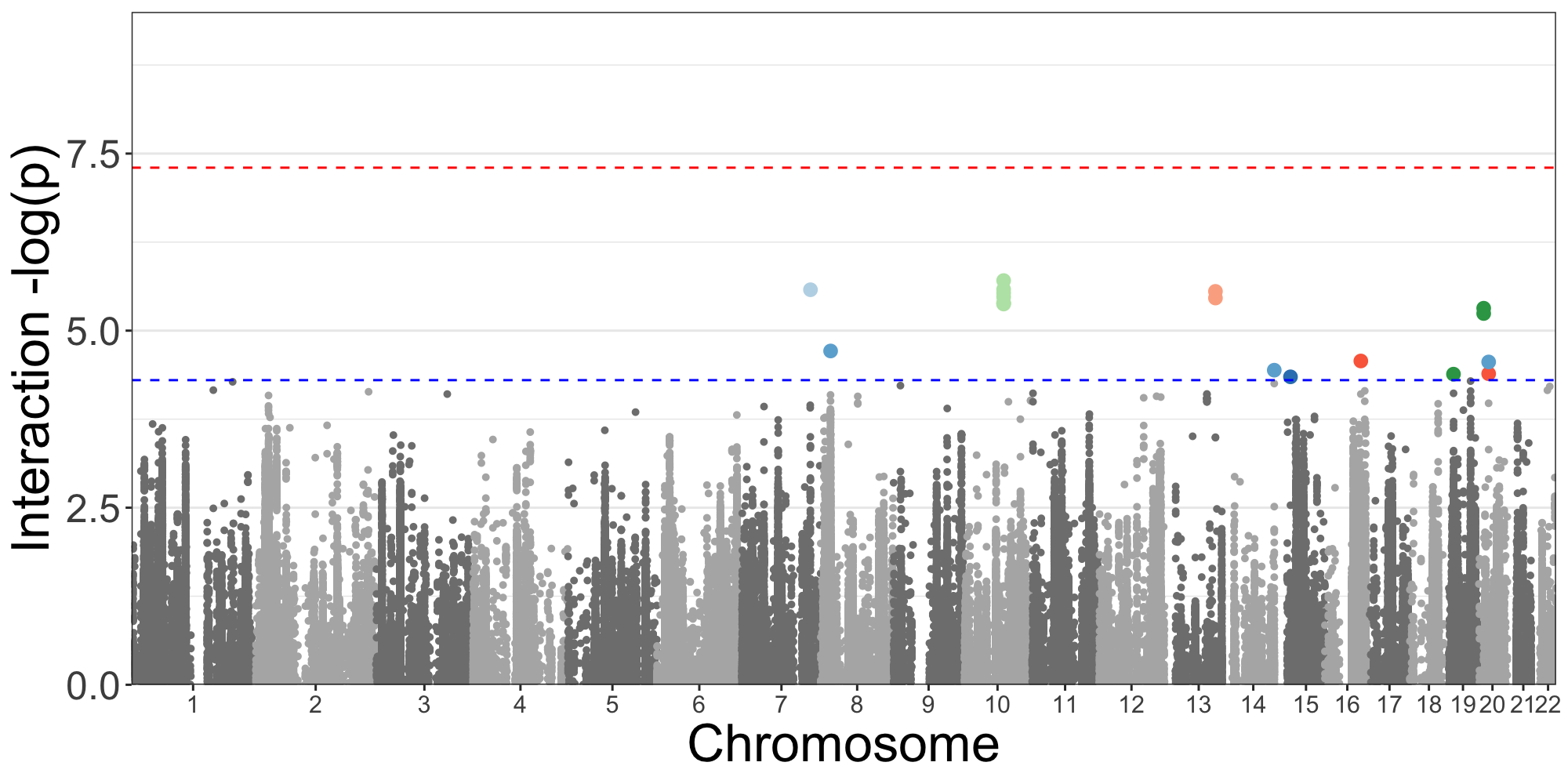

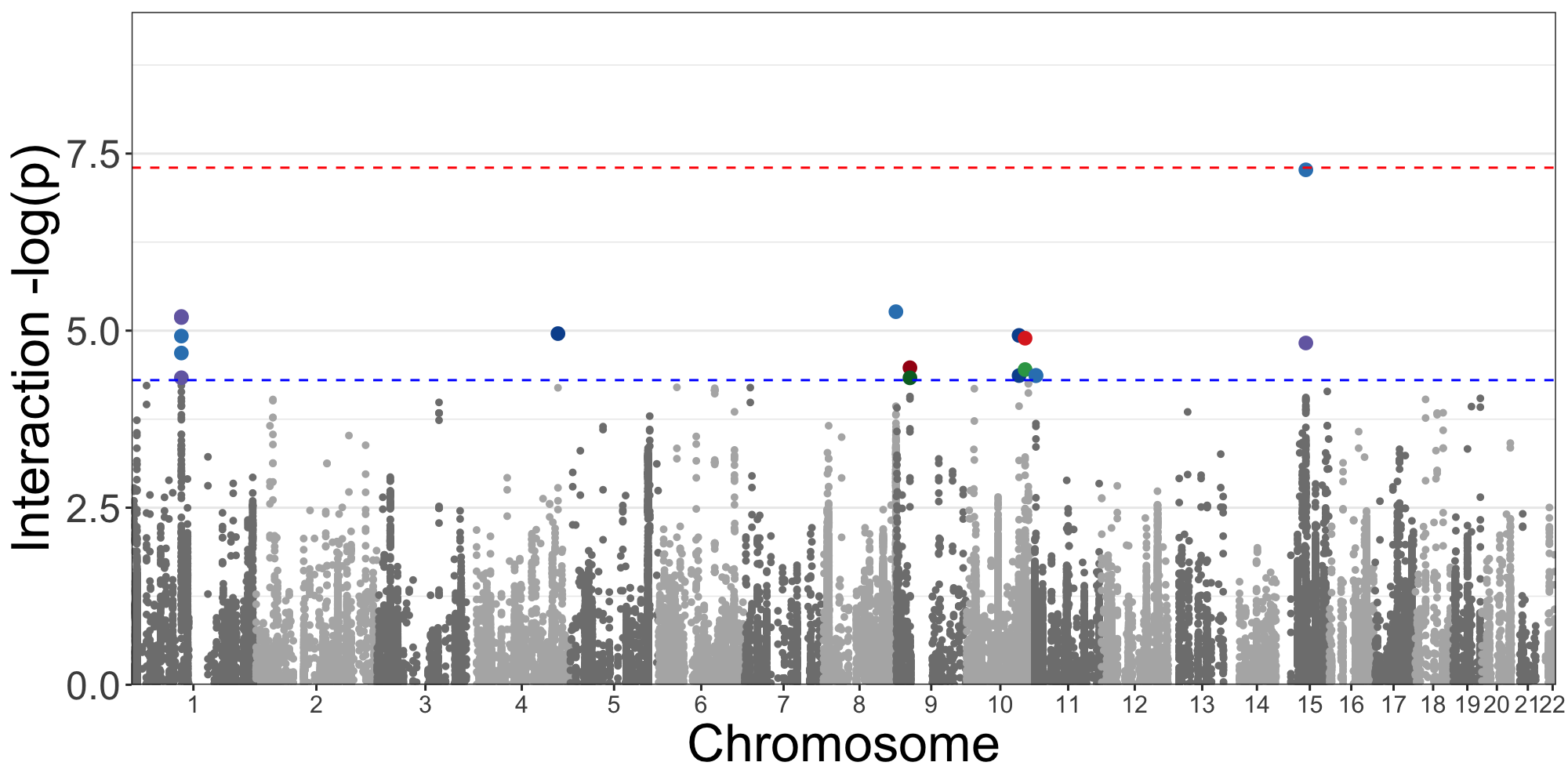

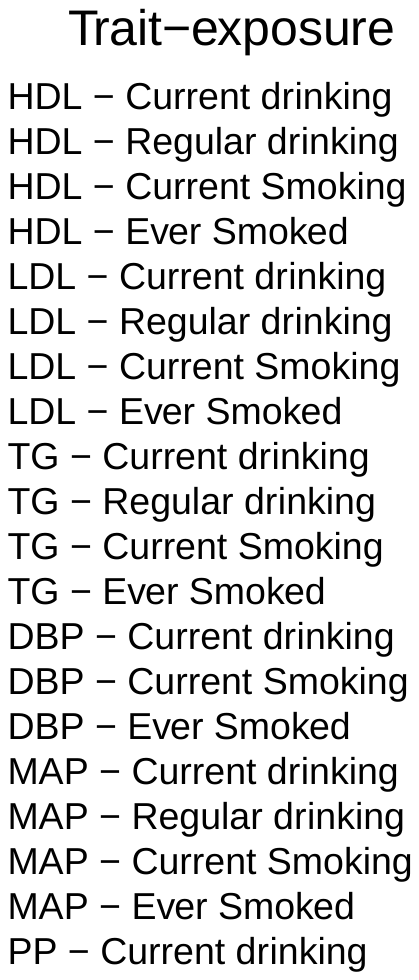

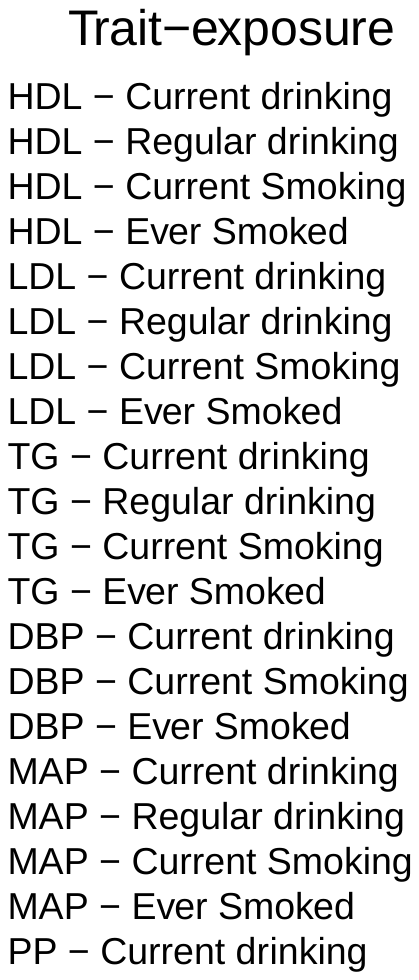

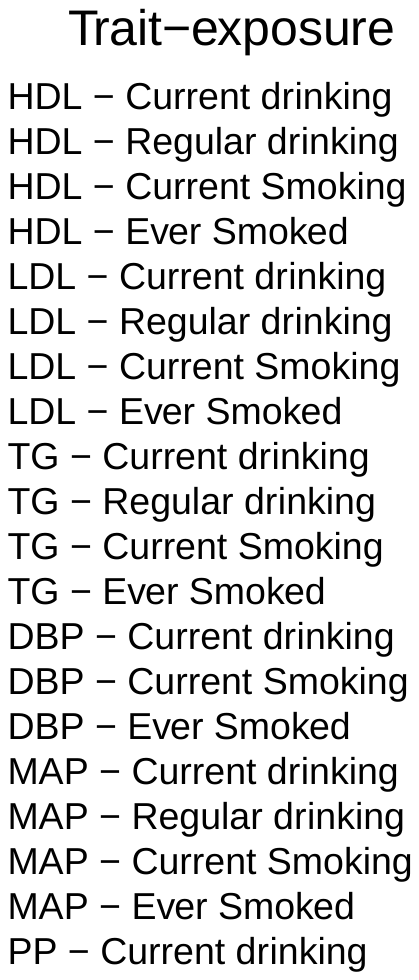

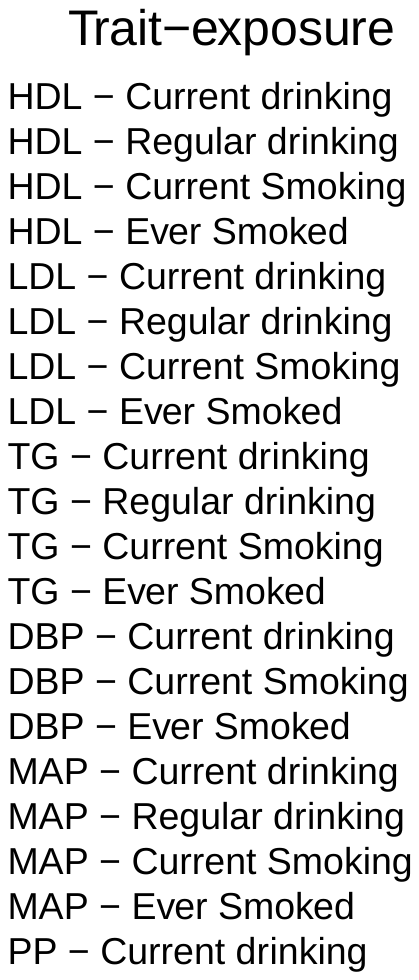

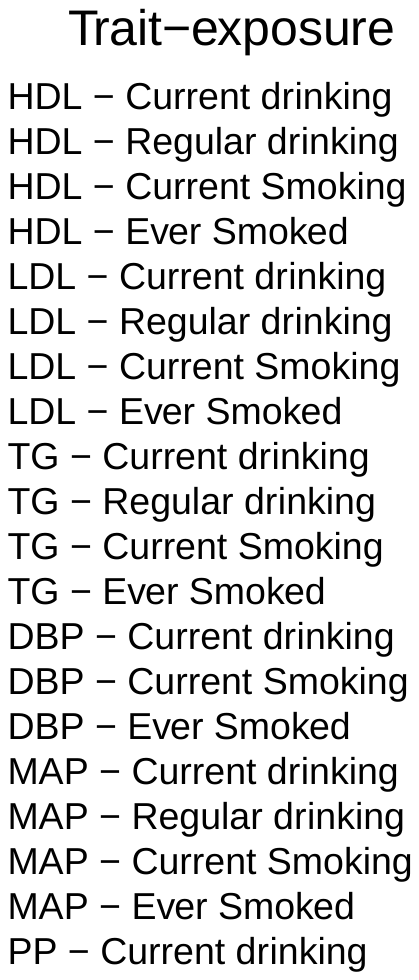

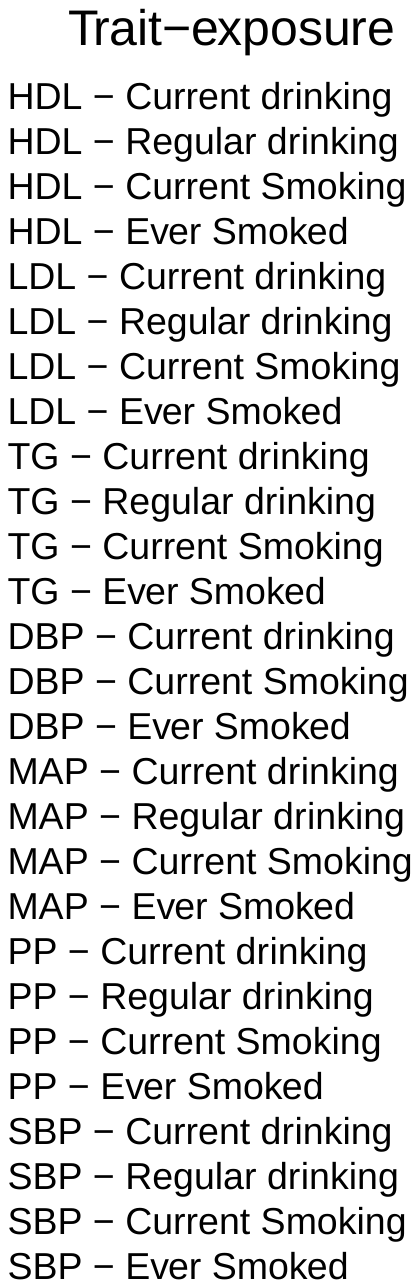

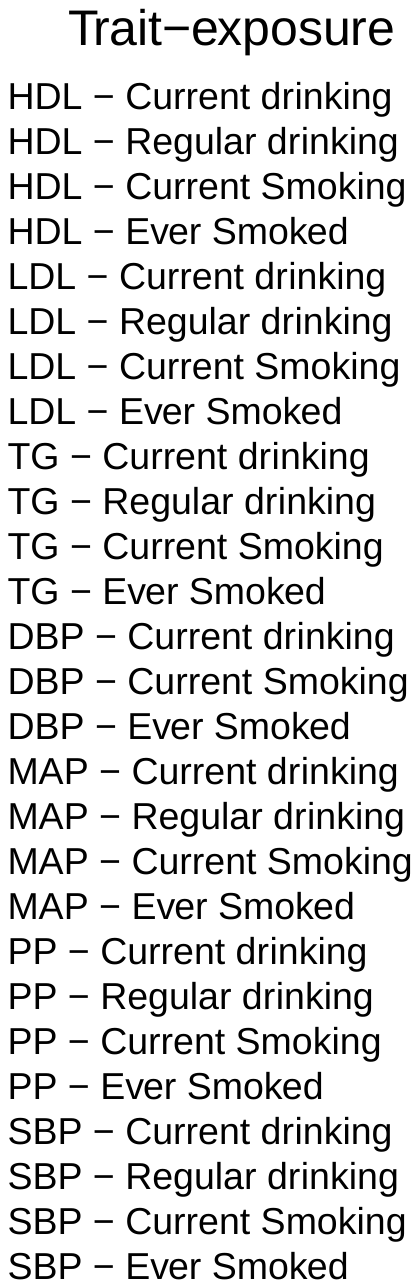

**Supplemental Figure 2.** (A) Regional Plot *GCNT4* × Current Smoking and HDL (African Ancestry); (B) Summary of Relevant Data

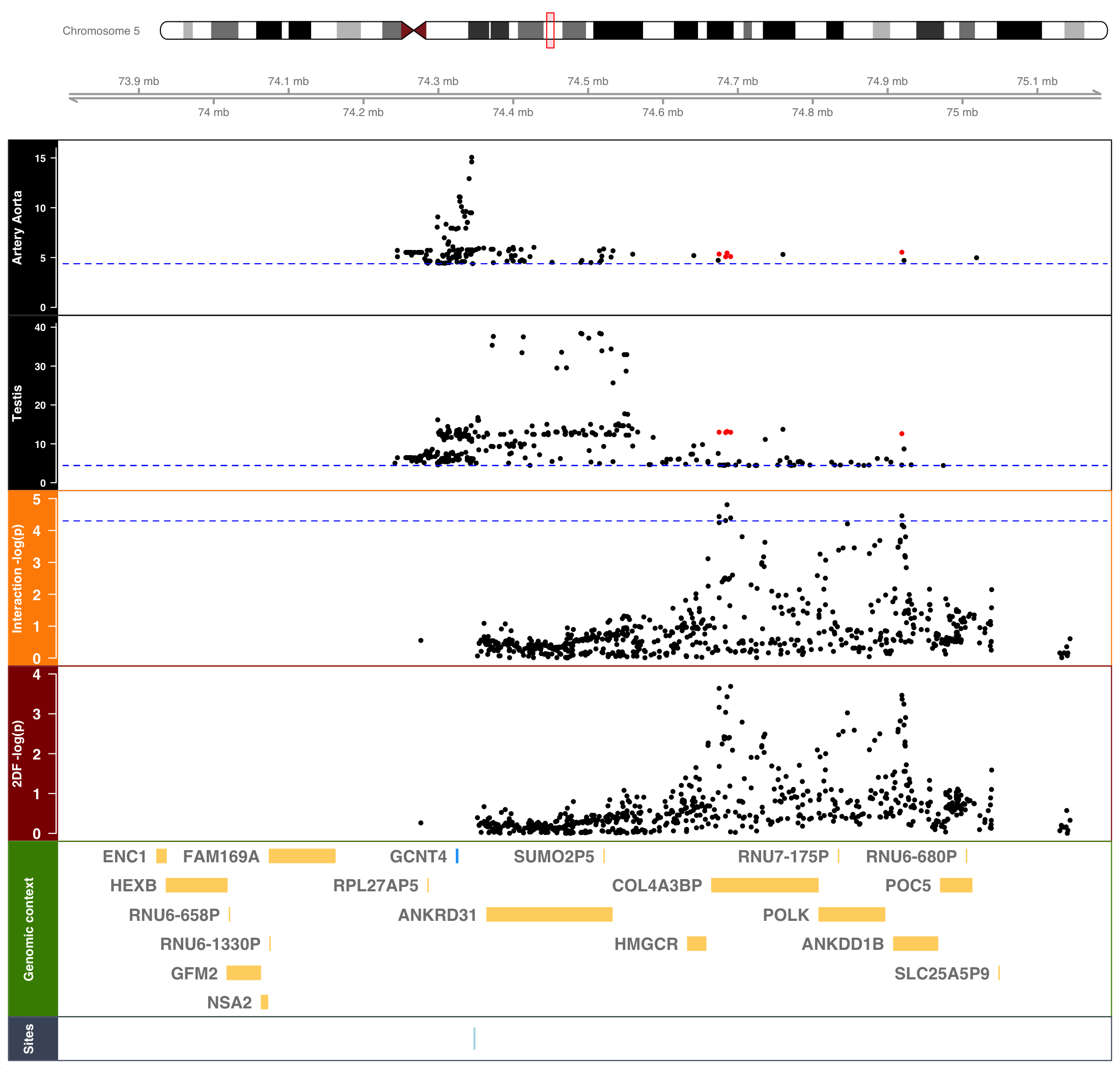

A.

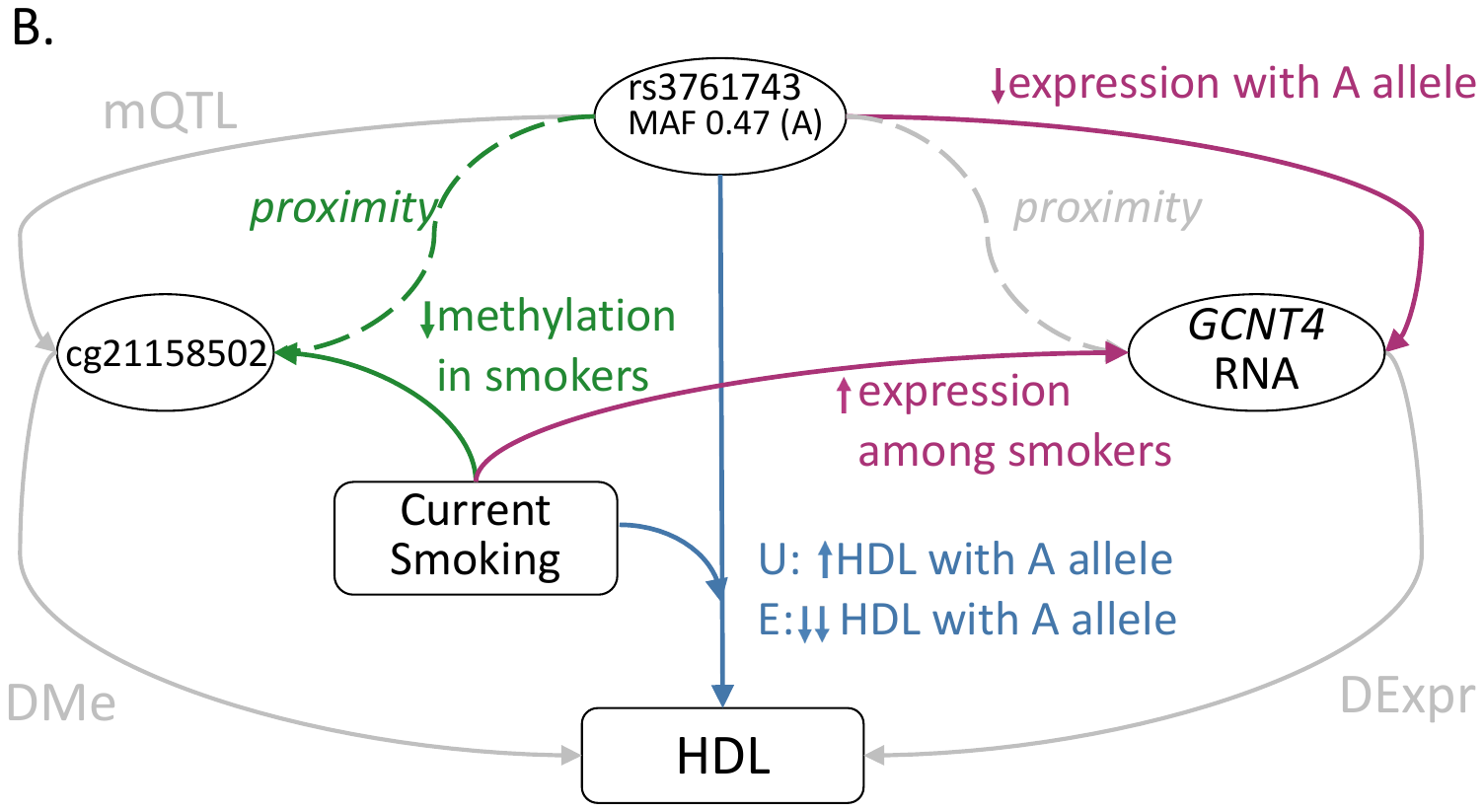

**Supplemental Figure 3.** (A) Regional Plot *PTPRZ1* × Ever Smoked and HDL (African Ancestry); (B) Summary of Relevant Data

A.

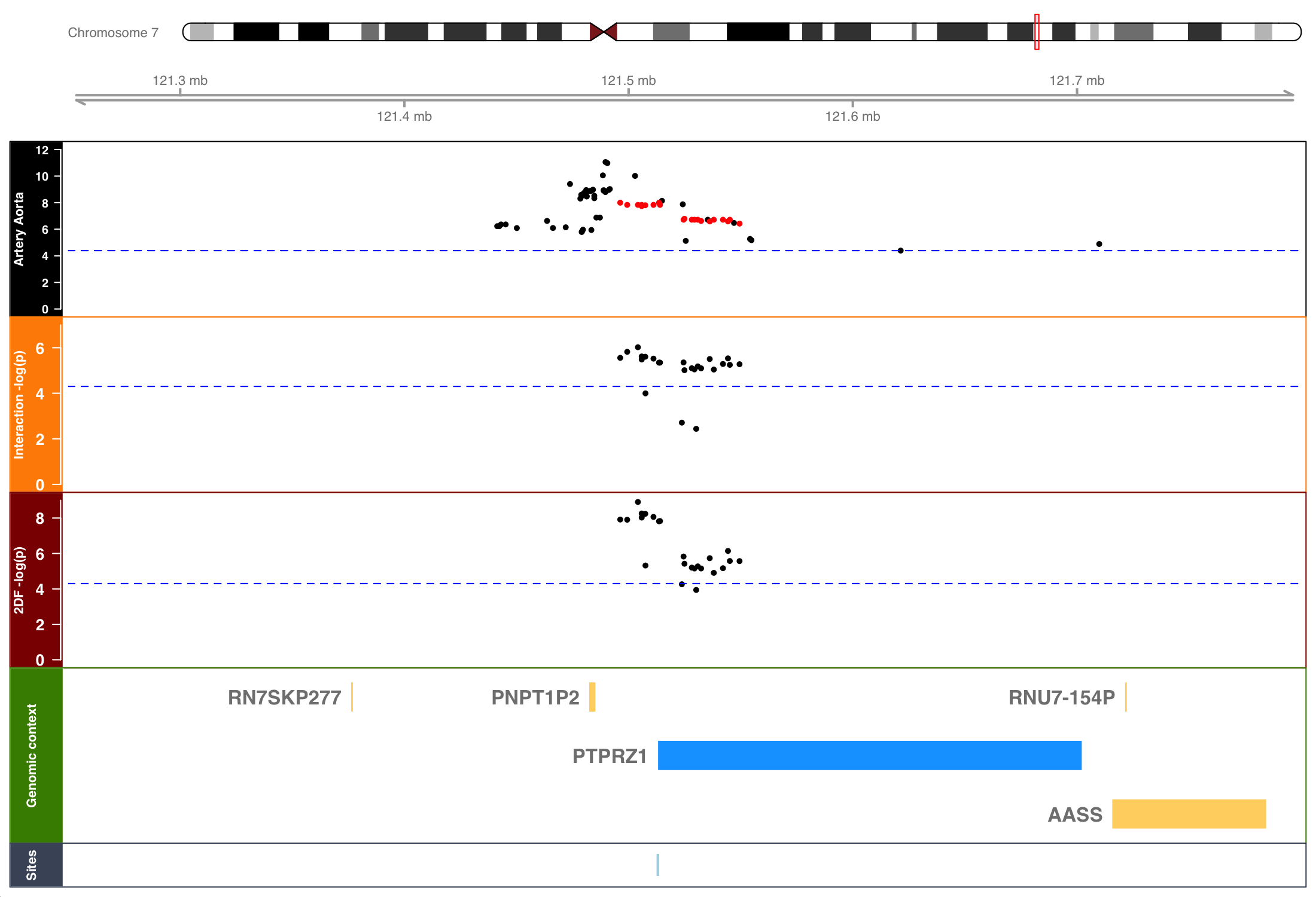

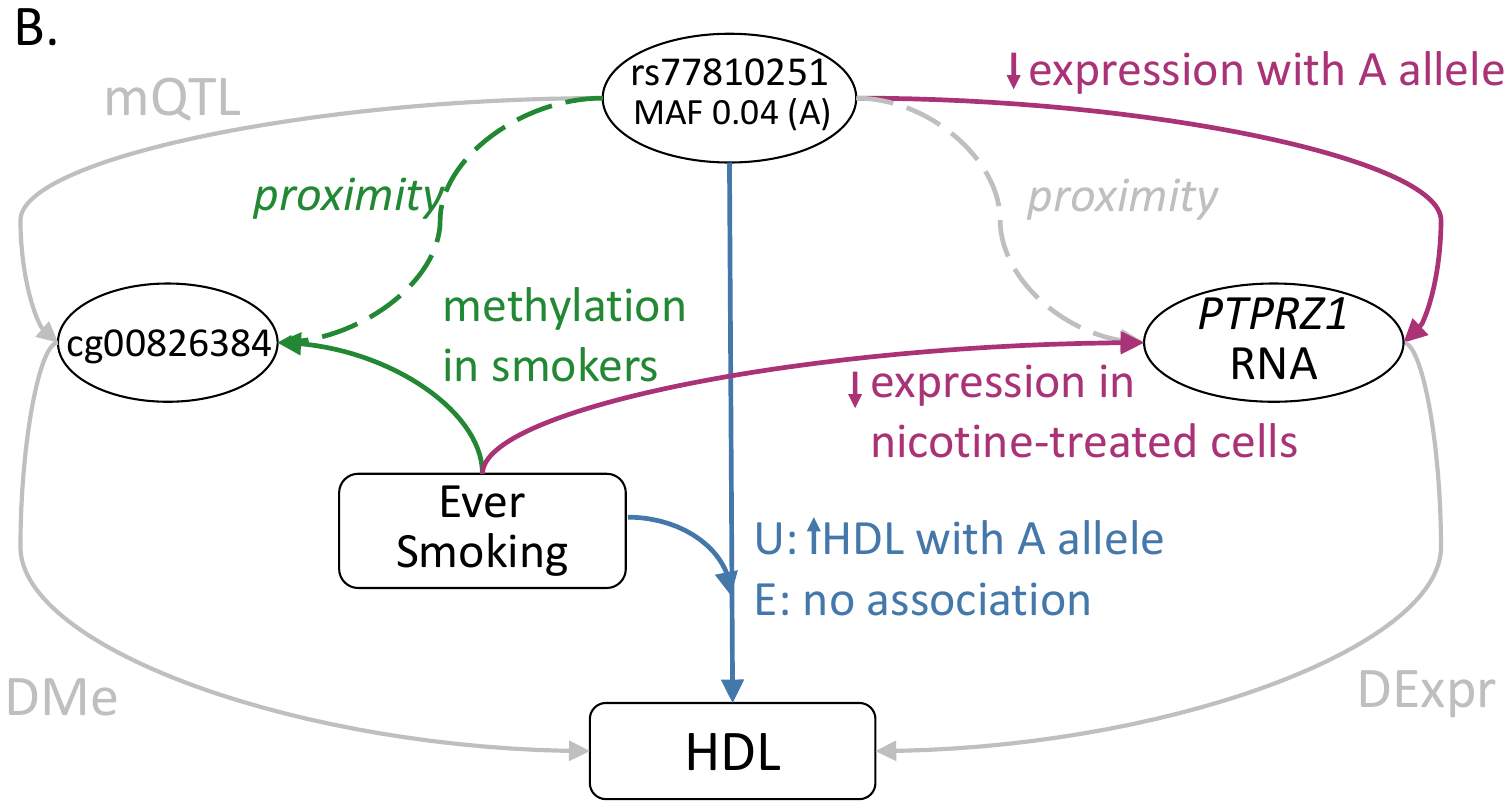

**Supplemental Figure 4**. (A) Regional Plot *SYN2* × Current Smoking on PP (African Ancestry); (B) Summary of Relevant Data

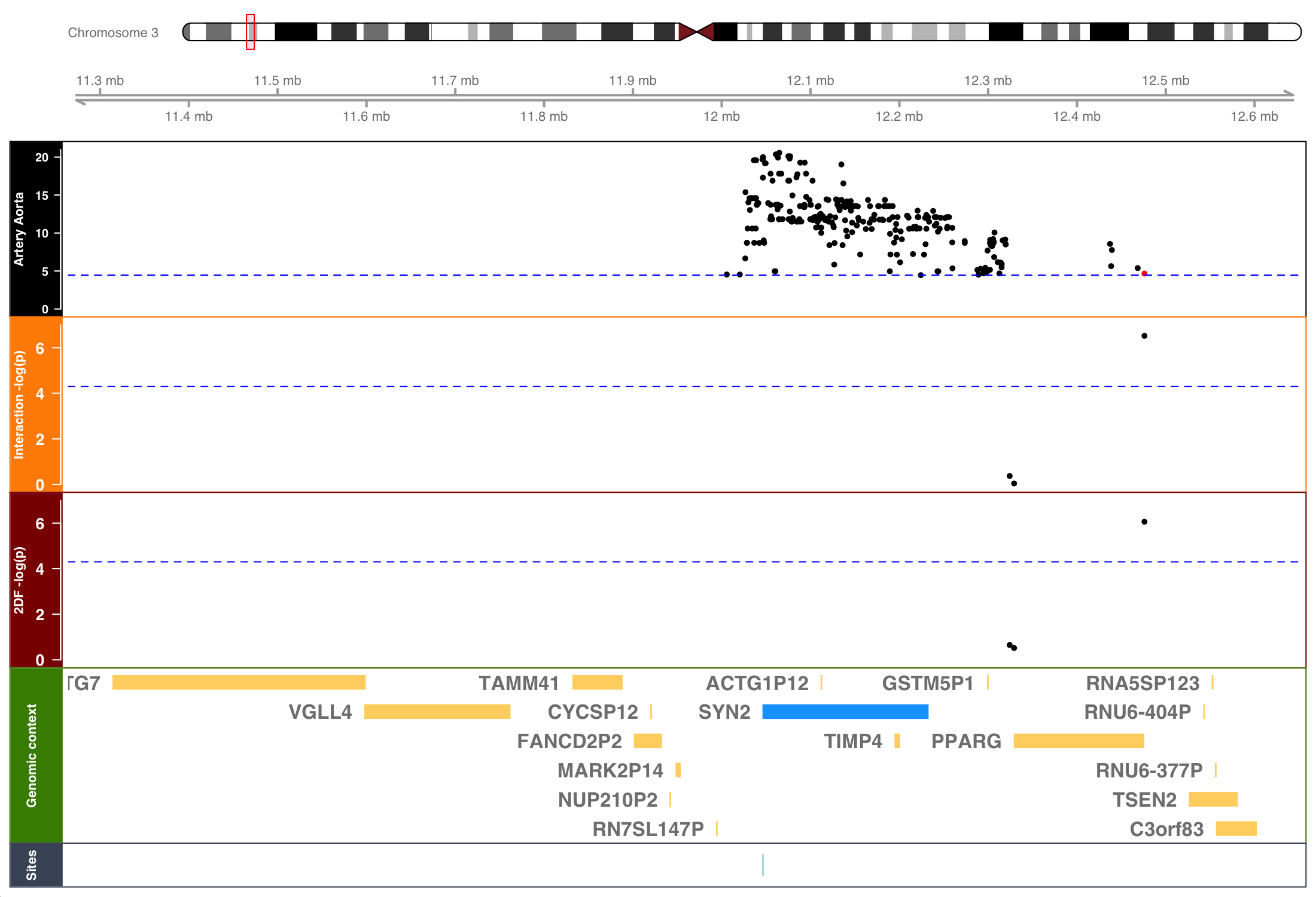

A.

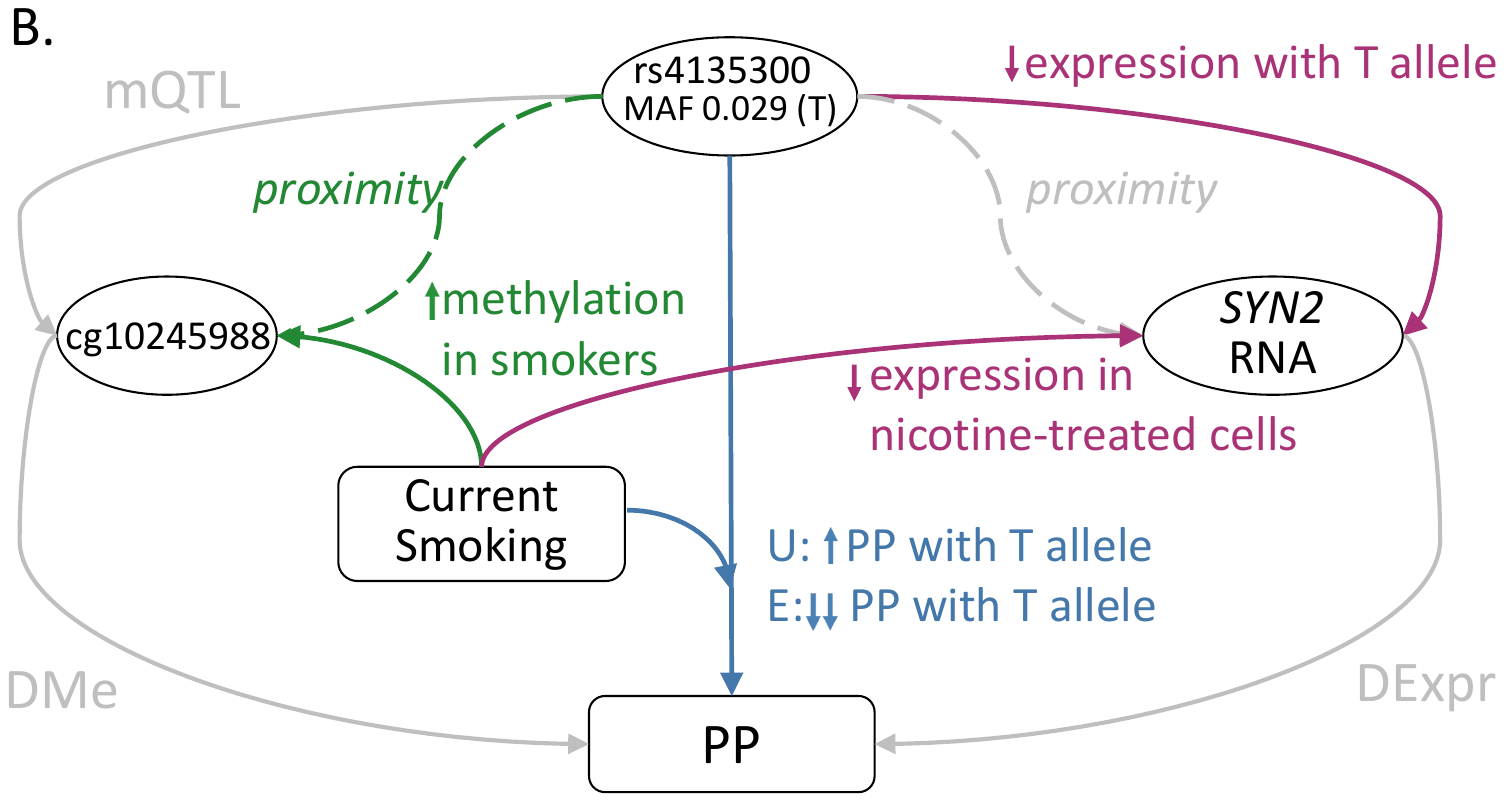

**Supplemental Figure 5**. (A) Regional Plot *ALDH2* × Ever Smoked and MAP (Asian Ancestry); (B) Summary of Relevant Data

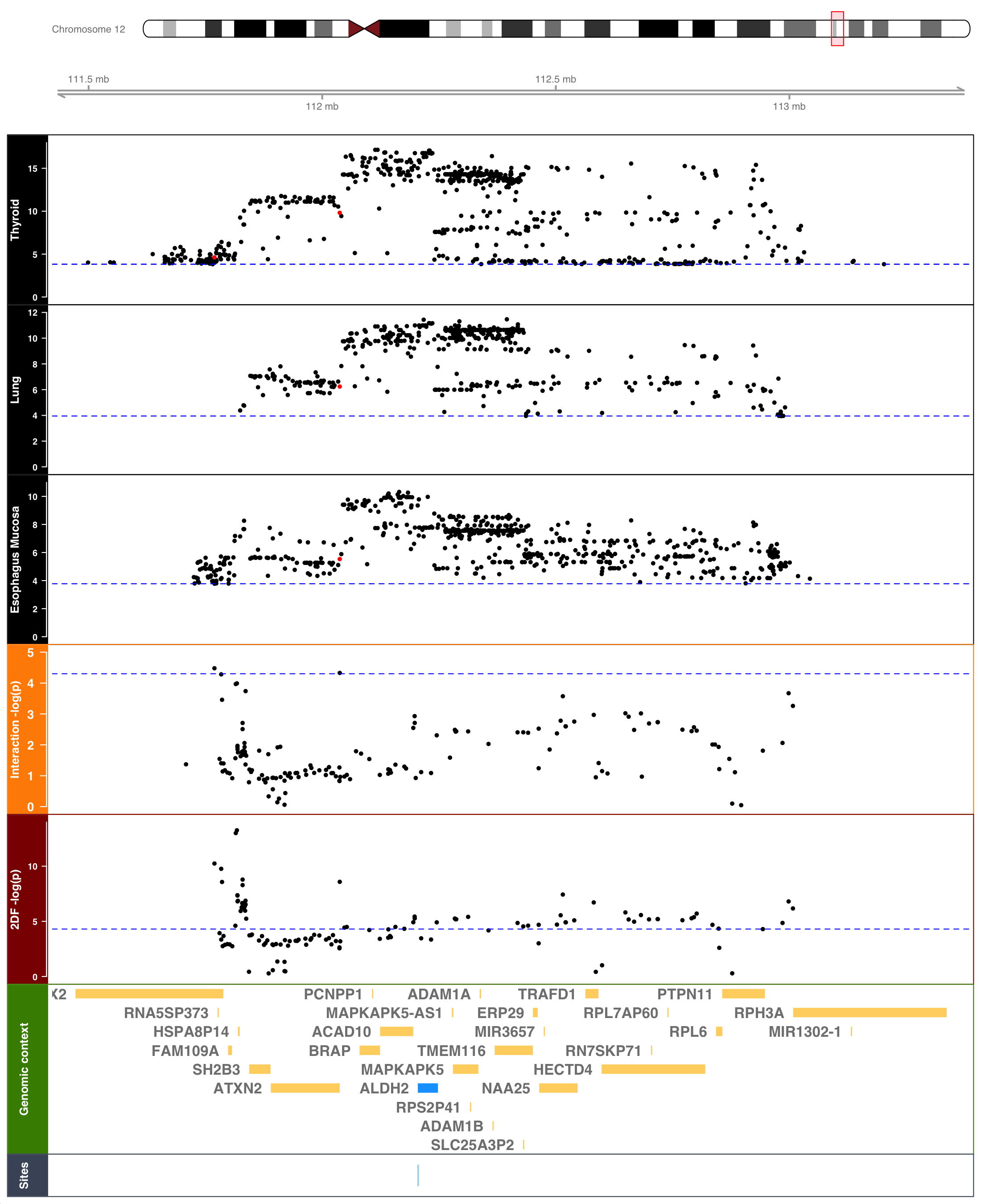

A.

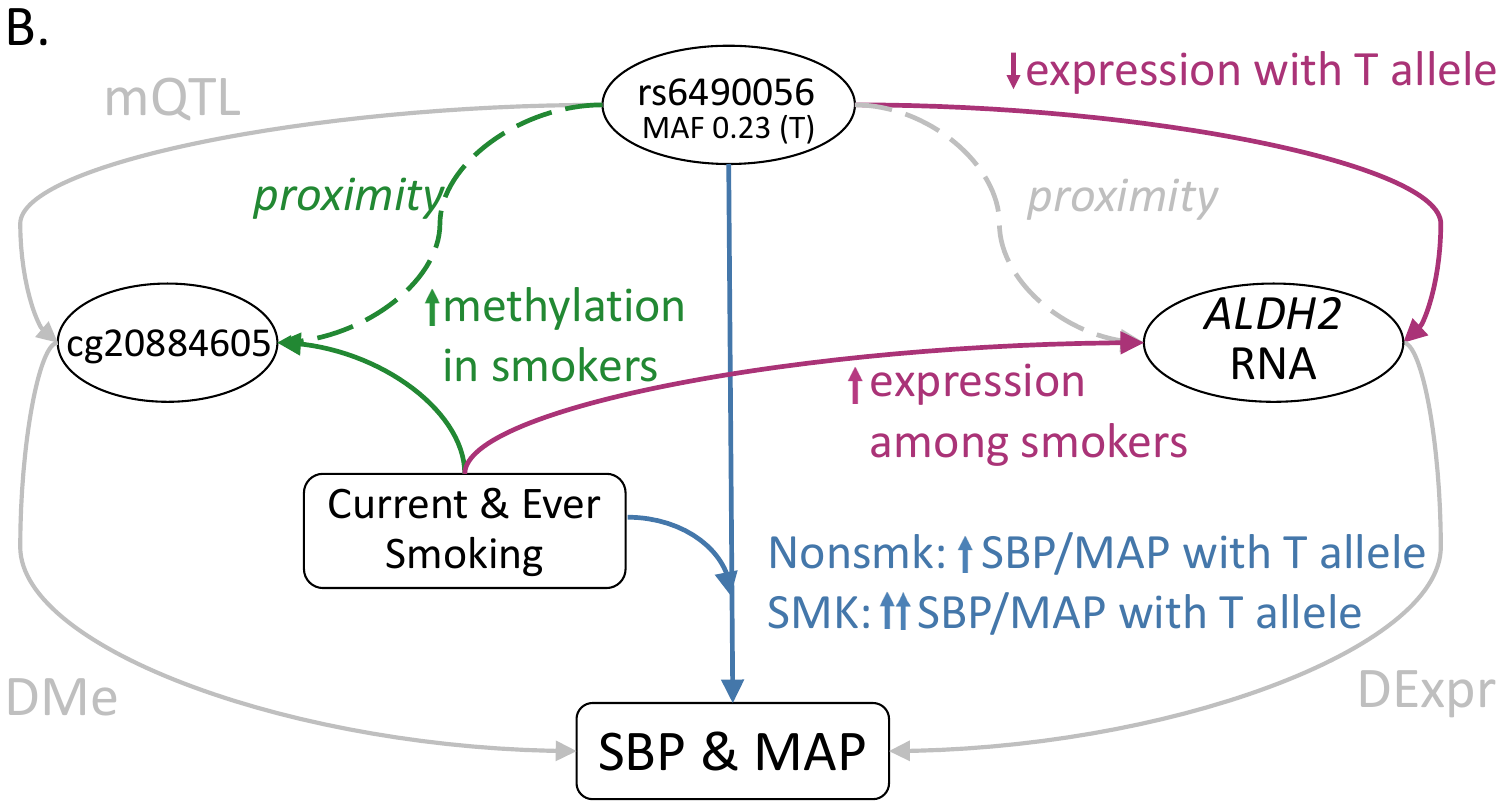

**Supplemental Figure 6.** Regional Plot *ALDH2* × Current Smoking and SBP (Asian Ancestry)

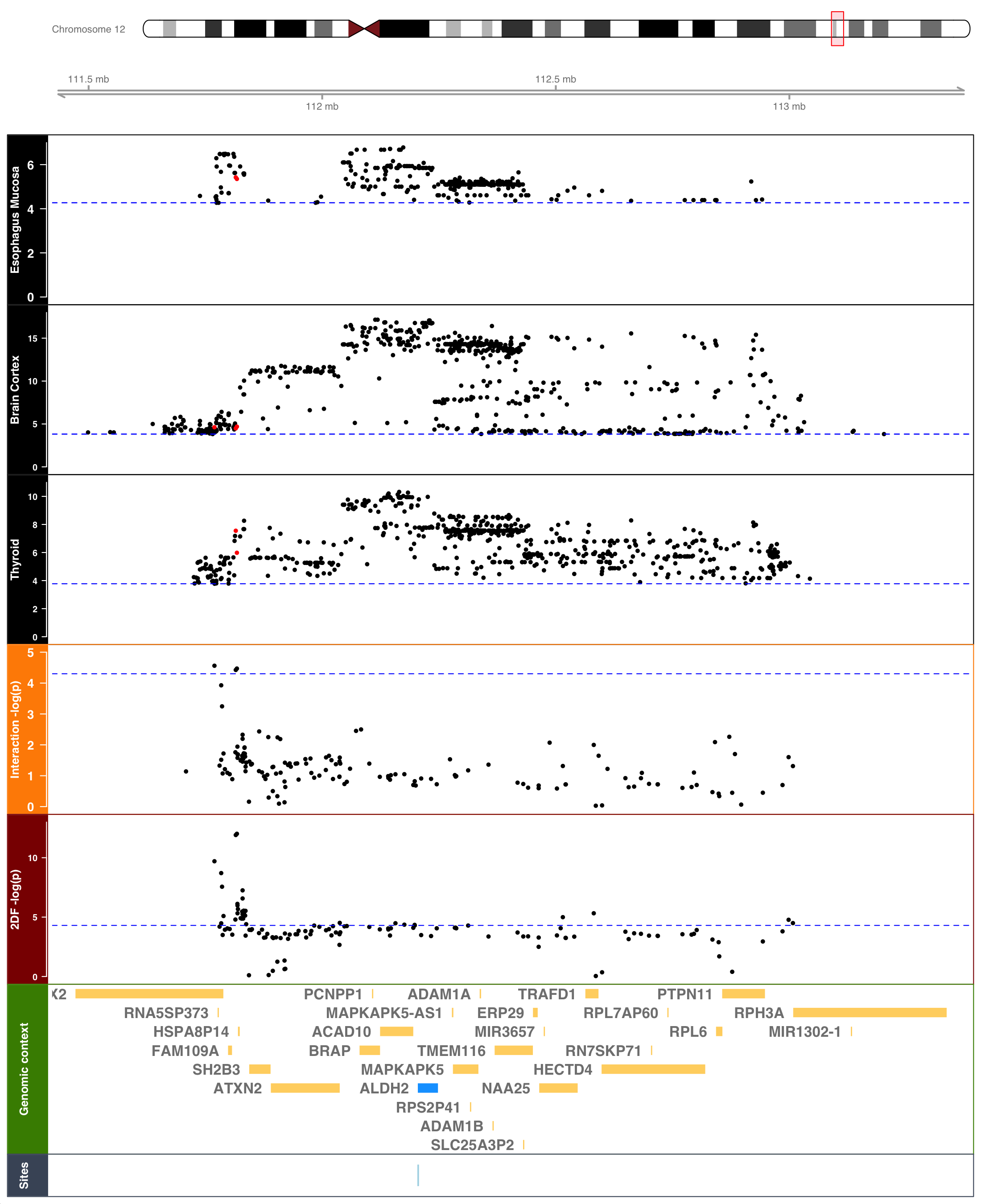

**Supplemental Figure 7.** Regional Plot *ALDH2* × Ever Smoked and SBP (Asian Ancestry)

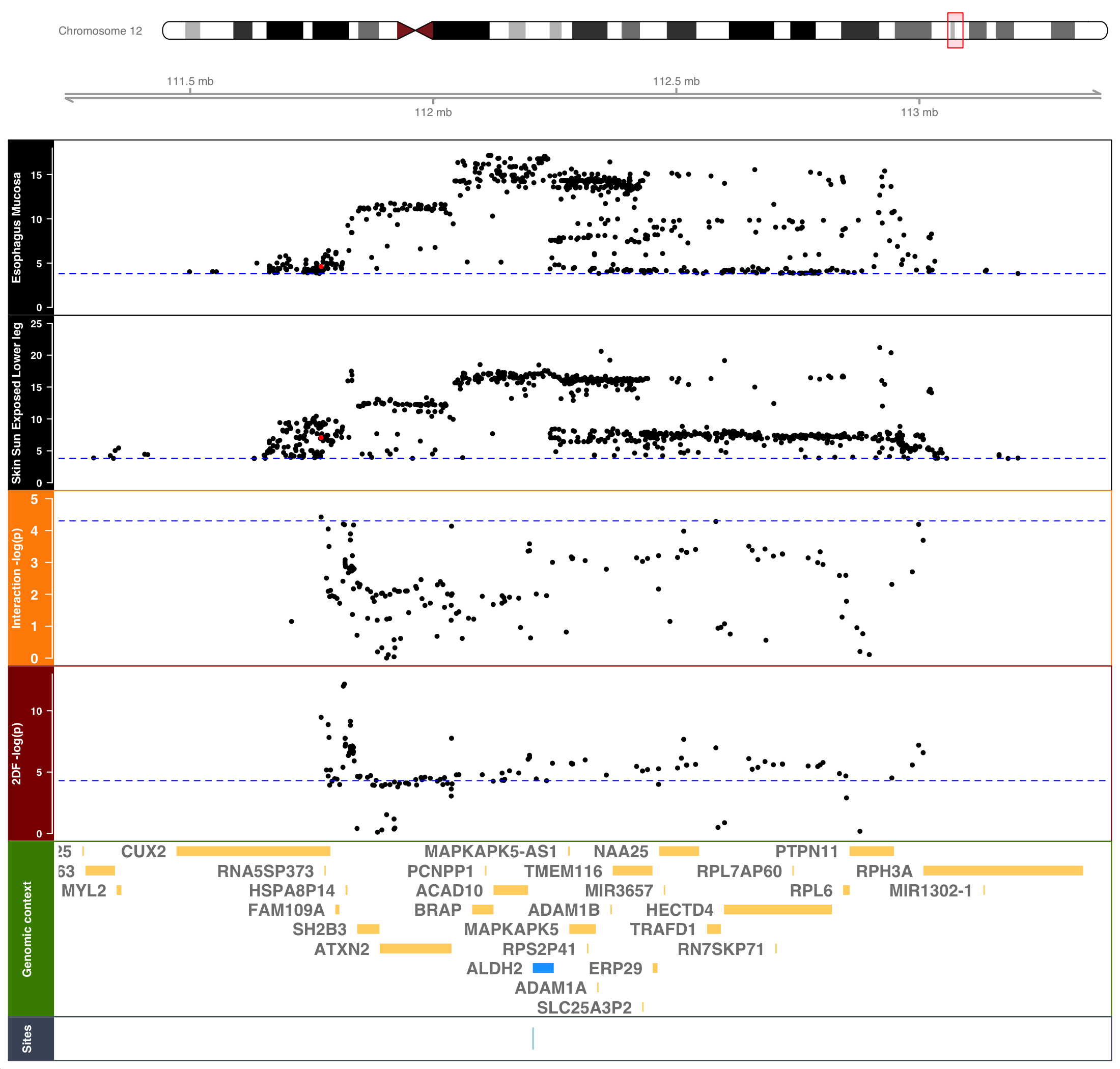

**Supplemental Figure 8.** (A) Regional Plot *TMEM116* × Ever Smoked and MAP (Asian Ancestry); (B) Summary of Relevant Data

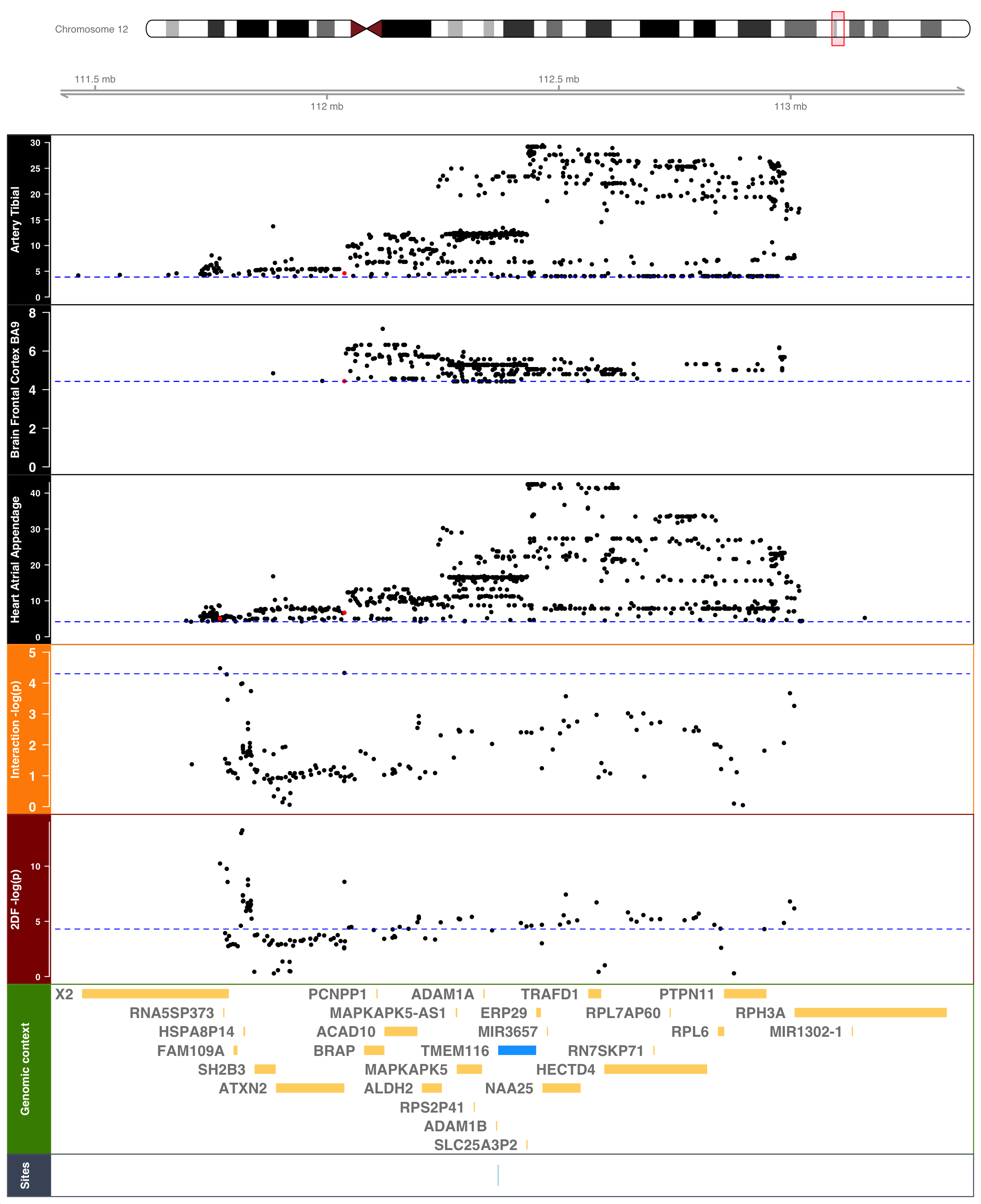

A.

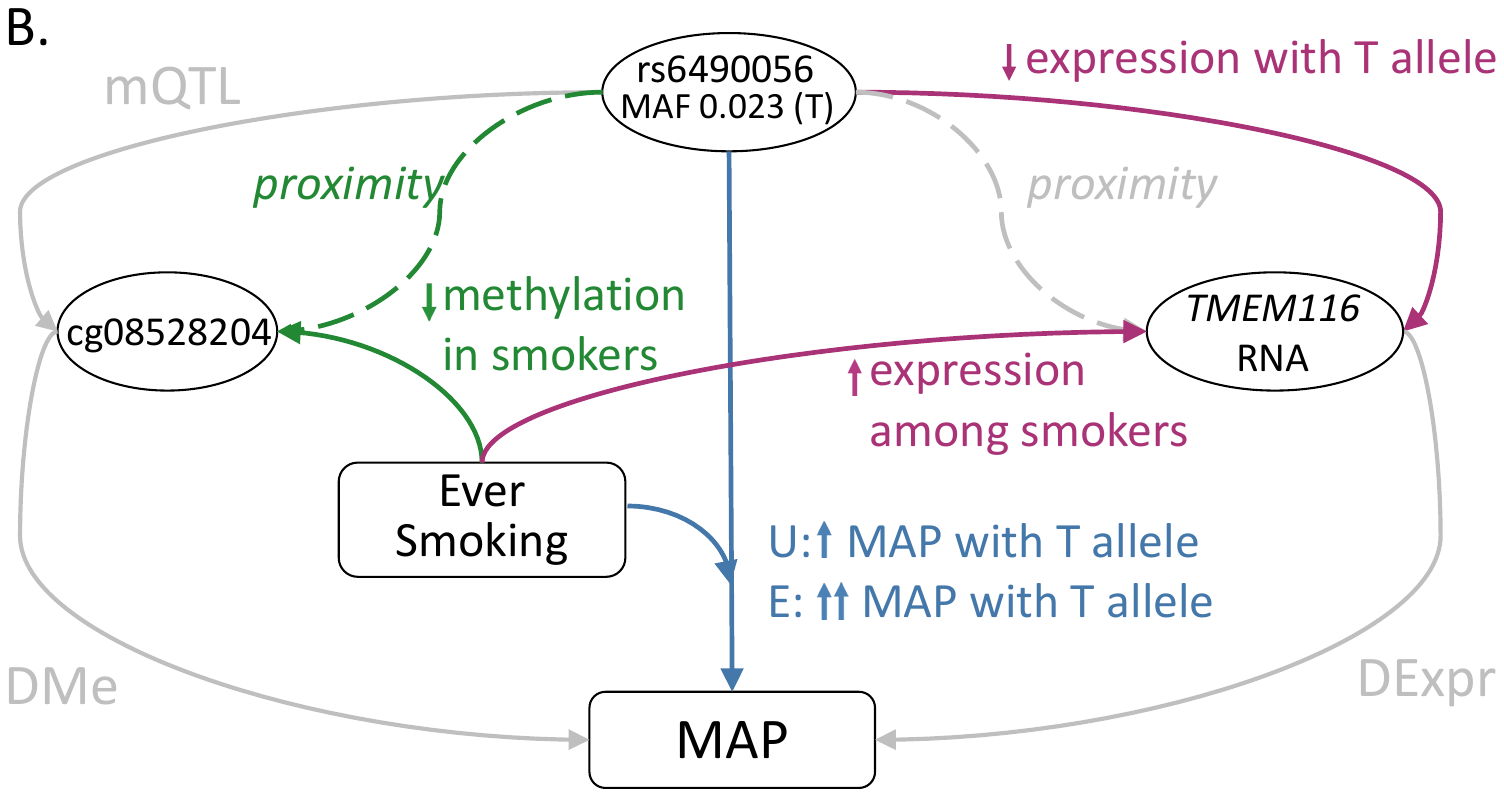
